# Mapping the causal chain from genetic risk variants to lipid dysmetabolism in Parkinson’s disease

**DOI:** 10.1101/2025.07.14.25331535

**Authors:** Ruth B. De-Paula, Jonggeol Kim, Herve Rhinn, Hiba Saade, Fatima Chavez, Téah Segura, Maria Valeria Lozano, Michelle Etoundi, Karla Silos, Viktoriya Korchina, Harshavardhan Doddapaneni, Eric Venner, Joseph C. Masdeu, Valory Pavlik, Melissa M. Yu, Chi-Ying R. Lin, Joseph Jankovic, Aron S. Buchman, Donna Muzny, Richard A. Gibbs, Sarah H. Elsea, Asa Abeliovich, Peter Lansbury, Nora Vanegas-Arroyave, Chad A. Shaw, Joshua M. Shulman

## Abstract

Risk variants in multiple genes, including *GBA1*, strongly implicate lipid metabolism in Parkinson’s disease (PD) onset and progression. We show that common PD risk variants at the *serine palmitoyltransferase small subunit B* (*SPTSSB*) locus, a key regulator of *de novo* sphingolipid biosynthesis, are associated with increased *SPTSSB* brain expression and elevated plasma ceramides. Additional analyses strongly support our hypothesis that a common *SPTSSB* causal variant is responsible for PD risk as well as the expression and metabolic quantitative trait loci. To systematically identify biomarker signatures that mediate PD risk and pathogenesis, we analyzed comprehensive metabolome profiles from blood plasma in 149 PD patients and 150 controls. Multiple sphingolipids and fatty acid derivatives were perturbed in PD, and we identified both unique and shared features with the Alzheimer’s disease metabolome. A PD acylcarnitine signature was further replicated in metabolic data from human postmortem brain tissue, when comparing those with or without preclinical Lewy body pathology. Integrated analysis of complementary proteomic profiles from the same autopsy cohort revealed dysregulation of mitochondrial processes dependent on acylcarnitines, including fatty acid beta-oxidation, the tricarboxylic acid cycle, and oxidative phosphorylation. Our results reveal a causal chain linking genetic variation to altered gene/protein expression, lipid dysmetabolism, and the manifestation of PD.

## Introduction

More than 100 susceptibility loci have been discovered for Parkinson’s disease (PD); however, a major gap exists to understand how implicated genetic risk variants lead to the development of alpha-synuclein pathology and initial clinical manifestations of the disease^1,2^. Loss-of-function variants in the *Glucocerebrosidase* (*GBA1*) gene are among the most common and potent PD genetic risk factors, conferring at least 5-fold increased risk for PD^3^. *GBA1* encodes the lysosomal enzyme responsible for breakdown of glucosylceramide, a key precursor for membrane sphingolipids^4^. Common or rare variants in other genes encoding lysosomal enzymes or cofactors regulating metabolism of ceramides and sphingolipids have similarly been linked to PD risk (e.g. *SCARB2*, *ARSA, SMPD1*, and *GALC*) (**Figure 1A**)^5–9^. Based on studies in animal and cellular models, disruption of lysosomal metabolism and the accumulation of undigested sphingolipid species may directly promote alpha-synuclein pathology^10–12^. Thus, perturbations in sphingolipid/ceramide metabolism are a promising candidate mechanism by which genetic variants may drive PD clinical and pathologic manifestations^13,14^.

**Figure 1.**
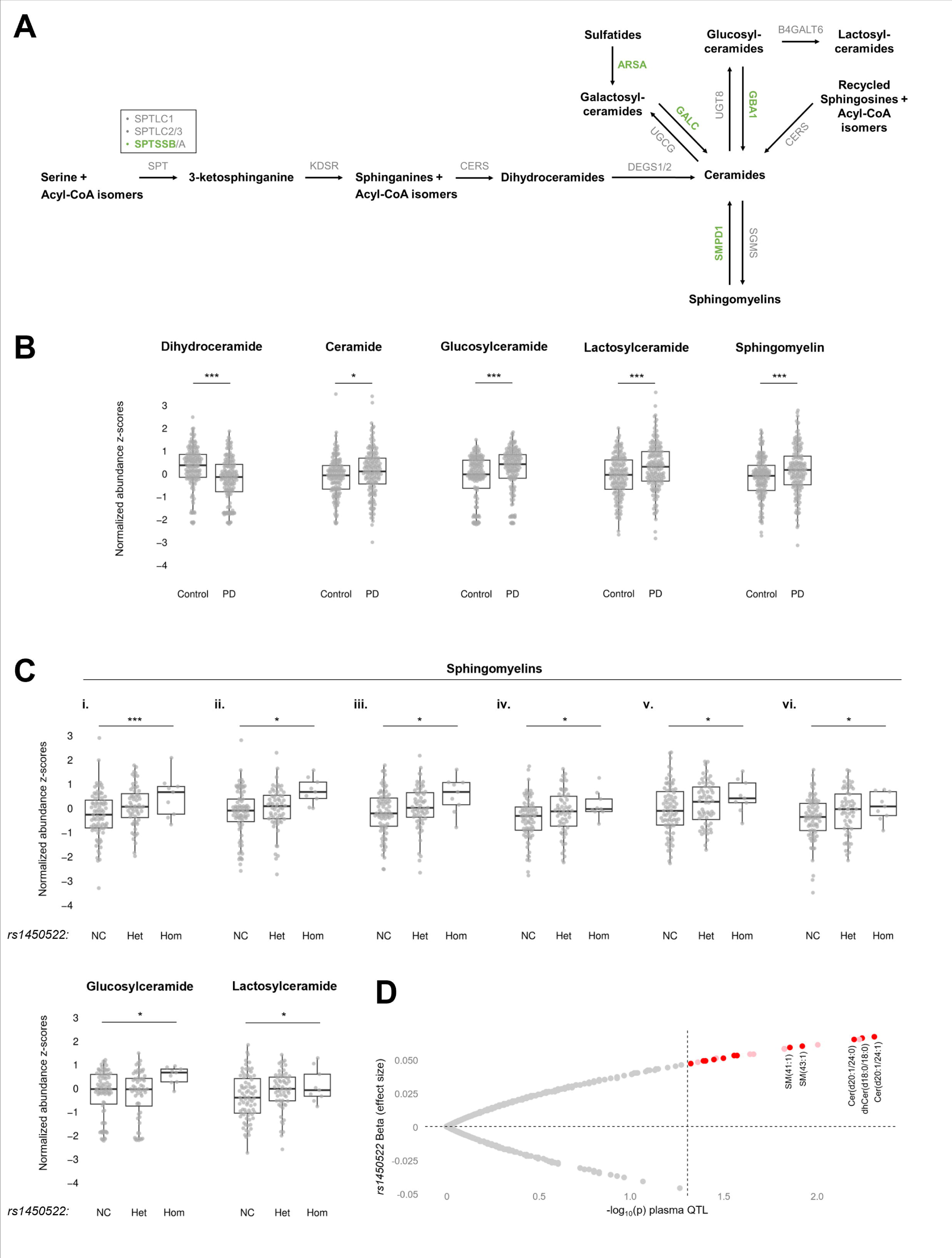
Sphingolipid perturbations in Parkinson’s disease (PD) cases and genetically at-risk controls. (A) Schematic showing the *de novo* sphingolipid synthesis and selected related metabolic pathways. PD risk genes are denoted in green. (B) Boxplots show representative sphingolipid perturbations in 149 PD cases versus 150 controls. Normalized residuals (z-scores) are plotted, following adjustment for age and sex. Significance testing was based on the likelihood ratio test. The following sphingolipids are shown: Dihydroceramide = N-stearoyl-sphinganine (d18:0/18:0), Ceramide = ceramide (d18:2/24:1, d18:1/24:2), Glucosylceramide = glycosyl ceramide (d18:1/23:1, d17:1/24:1), Lactosylceramide = lactosyl-N-nervonoyl-sphingosine (d18:1/24:1), Sphingomyelin = sphingomyelin (d18:2/24:1, d18:1/24:2). (C) Among 150 controls without PD, boxplots show sphingolipid increases in heterozygous (Het) or homozygous (Hom) carriers of the PD risk variant, *rs1450522*, at the *SPTSSB* locus when compared with non-carriers (NC). The whiskers denote the interquartile range between the first and third quartiles. Normalized residuals (z-scores) are plotted, following adjustment for age and sex. Significance testing was based on logistic regression considering an additive inheritance model, except for glucosylceramide, where a recessive model was employed. The following sphingomyelins are shown: (i) sphingomyelin (d18:1/22:1, d18:2/22:0, d16:1/24:1), (ii) sphingomyelin (d18:2/23:0, d18:1/23:1, d17:1/24:1), (iii) sphingomyelin (d18:2/21:0, d16:2/23:0), (iv) sphingomyelin (d18:2/24:1, d18:1/24:2), (v) sphingomyelin (d18:1/20:0, d16:1/22:0), (vi) sphingomyelin (d18:2/24:2). Glucosylceramide = glycosyl ceramide (d18:1/23:1, d17:1/24:1), Lactosylceramide = lactosyl-N-nervonoyl-sphingosine (d18:1/24:1). For complementary analyses in PD cases, see Supplemental Table 3 and Figure S3. (D) Volcano plot showing lipid metabolites associated with *rs1450522* meeting the suggestive significance threshold (unadjusted *p <* 0.05 dashed horizontal line), with sphingolipids highlighted in red. A total of 629 lipid species were interrogated in *n* = 4,492 individuals without known neurological disease ^51^. *, *p* < 0.05; ***, *p* < 0.005.

Prior to the appearance of cardinal motor signs, individuals with PD progress through preclinical and prodromal phases characterized by absent or minimal symptoms in association with brain pathology, making the discovery of sensitive biomarkers for early detection and monitoring of disease progression a priority^15^. Published studies in blood, CSF, and postmortem brain tissue suggest perturbations in sphingolipids as well as other lipid metabolites—or changes in the activity of enzymes like Glucocerebrosidase—may accompany PD^16–23^. However, interpretation of these studies to infer early or proximal, causal changes is challenging since most studies do not control for disease duration, and sphingolipid perturbations may also be a downstream consequence of PD pathophysiology.

Indeed, a preponderance of experimental data demonstrate that alpha-synuclein pathology can feedback to disrupt lysosomal function and lipid metabolism^11,12^. Beyond the lysosome, mitochondrial dysfunction has also been implicated in PD pathogenesis, and mitochondria are a critical hub for metabolism, participating in fatty acid catabolism and oxidative phosphorylation^1,24,25^. Integration of genetic and metabolic markers may be a promising strategy both to identify biomarkers and resolve causal pathways^26^. For example, in a recent large study integrating genotyping and cerebrospinal fluid metabolomics, common genetic variants in *arylsulfatase A* (*ARSA*) were associated with both PD risk and tyrosine derivatives from cerebrospinal fluid (CSF)^26^. Another challenge to robust PD biomarker discovery stems from the common co-occurrence of other age-related brain pathologies^27^, such as Alzheimer’s disease, which may interact with PD pathology and potentially cause related metabolic changes^28^.

We have comprehensively profiled metabolism in 398 individuals (149 PD cases, 99 AD cases, and 150 controls) and analyzed together with genetic data on *GBA1* and related lysosomal genes. We integrated these results with complementary analysis of available genetic, metabolic, and proteomic data from additional cohorts, including from blood (*n*=4,492; *n* = 19,994) and postmortem brain tissue (*n* = 429), identifying promising biomarkers of lipid dysmetabolism in PD / Lewy body pathology. Our results further highlight a causal chain between risk variants at the *SPTSSB* locus, encoding a subunit of serine palmitoyl transferase (SPT), which drive increased sphingolipid production and altered fatty acid metabolism, leading to elevated PD risk.

## Materials and methods

### Human Subjects

All studies of human subjects were conducted in accordance with the principles expressed in the Declaration of Helsinki. The institutional review boards at Baylor College of Medicine (BCM) and the Houston Methodist (HM) approved both the biospecimen collection and sequencing (H-50254; PRO00034371). Subjects were recruited from the Parkinson’s Disease Center and Movement Disorders Clinic and Alzheimer’s Disease and Memory Disorders Clinic BCM and from Nantz National Alzheimer Center at HM during routine clinic visits between 2022-2024. All patients with PD and AD were diagnosed by neurologists with subspecialty training in movement disorders or memory disorders, respectively. Unrelated controls were screened using a validated questionnaire to exclude individuals with potential PD symptoms or those with a first-degree family history of PD or dementia^48^. All participants agreed to provide a blood sample for a biorepository along with clinical and demographic data.

We also leveraged published clinical, pathologic, demographic, genome sequencing, and metabolic profile data from the Religious Orders Study and Rush Memory and Aging Project (ROSMAP; see also Data Availability, below). All ROSMAP participants enrolled without known dementia and agreed to detailed clinical evaluation and brain donation at death^29^. The ROSMAP studies were approved by the Institutional Review Board at Rush University Medical Center (ROS IRB# L91020181, MAP IRB# L86121802). Each participant signed an informed consent, Anatomic Gift Act, and an RADC Repository consent (IRB# L99032481) allowing data and biospecimens to be repurposed. For the analyses of differential metabolism and protein expression, Lewy body cases were classified based on alpha-synuclein positive Lewy body pathology in the substantia nigra, neocortex, and/or limbic area (variable: *dlbany*)^30^, and AD pathologic diagnosis was based on high or intermediate likelihood of AD according to NIA-Reagan scores (variable: *niareagansc*)^31,32^.

### Genome Sequencing

For BCM/HM samples, DNA was extracted from peripheral blood cells using the E.Z.N.A. SQ Blood DNA Kit (Omega Bio-tek Inc.) or from saliva samples using the Gentra Puregene Blood Kit (DNA purification from body fluid protocol) (QIAGEN). Approximately 3 µg of high-quality genomic DNA, at a concentration of 30–50 ng/µl in 70 µl of TE or EB buffer, was submitted to the BCM Human Genome Sequencing Center (HGSC). We followed published library preparation and sequencing protocols^33^. Libraries were prepared using KAPA Hyper PCR-free reagents on Beckman robotic workstations, with DNA shearing by Covaris, dual-size selection with AMPure XP beads, and ligation of Illumina dual-index adapters. Libraries were QC-checked by Fragment Analyzer and qPCR, then sequenced on the Illumina NovaSeq 6000 platform using S4 flow cells to generate 150 bp paired-end reads. BCM WGS data was aligned to hg38^34^ followed by variant calling using lllumina’s Dynamic Read Analysis for GENomics (DRAGEN) software, v3.7.8^35^. An in-house R pipeline was used to extract all single nucleotide variants (SNVs) within multi-sample Variant Call Files (VCFs). This pipeline generated a tabular output containing variant coordinates (chromosome number, start coordinate and end coordinate), reference and altered nucleotides, allele zygosity (homo/heterozygous), and slash-separated lists of subjects carrying each variant. For the *SPTSSB* analyses, we interrogated *rs1450522* (hg38 chr3:161,359,842 A>G), based on published PD genome-wide association studies^5^. For *GBA1*, we queried the well-established PD risk variants, L444P, N370S, T369M, and E326K^36,37^.

### Metabolomics

For both BCM/HM plasma and ROSMAP brain samples, global metabolomics (HD4 platform) mass-spectrometry intensity values (representing area under the curve (AUC) of mass-spectrometry peaks and proportional to metabolite abundance) were generated by Metabolon (https://www.metaboloninc.com/), including batch correction, according to previously published methods^28^. In total, 1,409 plasma metabolites were detected from the BCM/HM cohort, and 1,055 brain tissue metabolites were available from ROSMAP (dorsolateral prefrontal cortex). As in prior published studies^38^, metabolites with missing values for 30% or more samples were excluded, resulting in 1,171 metabolites (BCM/HM) and 685 metabolites (ROSMAP) for consideration in our analyses. The remaining missing values were imputed as minimum, and the data table was log2 transformed and mean centered (z-scores) for use in parametric statistical analyses.

#### Quantitative trait locus analysis

For single variant analyses, boxplots of metabolite normalized residuals (corrected for age at sample collection and sex) were plotted using the R package *ggplot2* (v. 3.4.4)^39^. Additive and recessive linear models (corrected for age at sample collection and sex) were used to test the association of heterozygous and homozygous *SPTSSB* rs1450522 variants on metabolite levels.

Colocalization analysis was performed using the *coloc* package in R^40^ and hg19 genome build, adopting a published pipeline^6,41^. PD GWAS summary statistics^42^ were extracted for variants from the *SPTSSB* locus (+/- 500kb), along with available eQTL / mQTL data (GTEx, BrainMeta^43^, and OmicScience^44,45^). Besides *SPTSSB*, secondary analyses considered eQTLs for other genes at the locus (*PPML1*, *B3GALNT1*, *NMD3* and *OTOL1*). We considered posterior probabilities (PP), including PP.H3, representing the probability that both traits (PD phenotype and QTL) are associated but have independent causal variants, and PP.H4, denoting the probability that both traits share a common causal variant. Co-localization was considered significant if PP.H4 > 0.5 and PP.H4 > PP.H3.

For those genes and traits showing significant co-localization, we performed Summary-data-based Mendelian Randomization (SMR), using published methods^43^. We used the same QTL and PD GWAS summary statistics (also hg19) as in the co-localization analyses detailed above. Genetic instruments were constructed using only GWAS significant variants (*p* < 5x10^-8^). We did not perform clumping since all variants were in high LD. Each mQTL was annotated to the gene most closely localized to the variant. SMR and HEterogeneity In Dependent Instruments (HEIDI) horizontal pleiotropy analysis was performed in a Linux-based 64 bits computer cluster, using SMR version 1.3.1. eQTL formatted input files (BESD/ESI/EPI) were obtained online (https://yanglab.westlake.edu.cn/software/smr/#DataResource). mQTL summary statistics were converted to BESD format using *--matrix-eqtl-format* and *--make-besd* flags. ESI files were annotated to include chromosome, position, effect allele, other allele, and effect allele frequency. EPI files were similarly annotated to include chromosome, probe name, central gene position, gene name, and gene strand for all genes within the *SPTSSB* LD locus. Finally, SMR and HEIDI tests were run using *--bfile* for defining PLINK 1000 Genomes European reference binaries; *--beqtl-summary* and *--gwas-summary* flags for defining QTL and GWAS inputs, respectively; *--peqtl-smr 0.05* and *--peqtl-heidi 0.05* for defining statistically significant QTLs; and *--smr-multi* for defining a multi-SNP analysis.

#### Analysis of differential metabolites

The likelihood-ratio test (LRT) was used to assess dysregulation of metabolites in plasma (clinical PD) or brain tissue (Lewy body pathology). Linear regression models (full vs. reduced) were constructed for each metabolite and used to calculate LRT statistics using R^46^. The full (complete) model, included contrasts and covariates; whereas the reduced model included covariates only, as below.

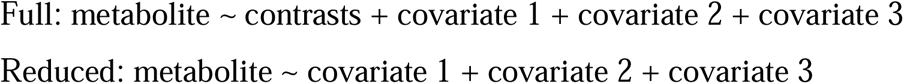

Compared with standard linear regression, the LRT approach allows for increased flexibility and improved fit for comparing complex, nested models. Analyses of plasma metabolites (BCM/HM cohort) included covariates for sex and age at sample collection; whereas analyses of brain tissue metabolites (ROSMAP) included covariates for sex, postmortem interval (PMI), and age of death. Residuals of reduced models were used for plotting and performing downstream analyses.

Volcano plots were generated using the R package *ggplot2* (v. 3.4.4)^39^. Venn diagram overlaps were calculated using *nVenn* (https://degradome.uniovi.es/cgi-bin/nVenn/nvenn.cgi), and we performed the Fisher test to assess statistical significance of the observed overlaps. The background of metabolites/proteins used to calculate overlap significance was the number of all molecules used for the differential expression analysis.

#### Proteomics

ROSMAP proteomics data based on untargeted, tandem mass tag mass spectrometry from dorsolateral prefrontal cortex tissue was downloaded from the AD Knowledge Portal (syn21448334)^47^. We excluded peptides with missing values for 30% or more samples, the remaining missing values were imputed as minimum, and peptide abundances mapping to the same protein were summed, resulting in abundances for 7,596 proteins. Data were log2 transformed and mean-centered (z-scores) prior to parametric statistical analyses. For differential protein expression, we followed the same LRT methods as for the analysis of differential metabolism (see above).

Overrepresentation analysis (ORA) of differentially expressed gene sets was implemented in MetaScape (https://metascape.org/gp/index.html)^48^. Background was set for all 6,511 proteins detected from ROSMAP cortical proteomics. A false discovery rate (FDR) < 0.05 was used as the significance threshold. Representative mitochondrial-related genes and ontology terms were queried for graphical display.

#### Statistics

Given the exploratory nature of our analyses, we considered an unadjusted *p* < 0.05 as meeting a suggestive level of significance, worthy of follow-up and further consideration. For the metabolome-wide analysis from either plasma or brain and proteomics analysis from brains, we additionally computed a multiple hypothesis adjusted p-value using the false discovery rate (FDR) method. For colocalization, we used PP.H4 > 0.8 to define robust co-localization. For SMR, we used SMR and SMR MultiSNP *p* < 0.05 to define SNP and locus significance, respectively, and HEIDI *p* > 0.05 to define absence of pleiotropy. For overrepresentation analysis using gene ontology terms, significance was set to an FDR-adjusted *p*-value < 0.05.

## Data availability

ROSMAP data is available for download from the Synapse AD Knowledge Portal (https://adknowledgeportal.synapse.org), including bulk brain tissue metabolomics (syn26007830)^28^, bulk and single-cell RNA-sequencing (syn3505720^29^, syn53366818^49^), proteomics (syn21449447)^47^, and whole genome sequencing (syn11707419)^50^. ROSMAP clinical, pathologic, and demographic data was also obtained from Synapse or requested directly from the Rush Alzheimer’s Disease Center. Data from the Surendran et al. metabolome-wide association analysis^44^ is accessible at (https://omicscience.org/apps/mgwas/mgwas.table.php); summary statistics for the *SPTSSB* locus were provided by Dr. Claudia Langenberg (University of Cambridge). We also downloaded summary statistics from the Cadby et al. lipidome-wide association analysis^51^. Complete summary statistics from all genetic and metabolomic data from the BCM-HM cohorts are included with the supplemental data as part of this work. Due to privacy concerns and the possible inadvertent release of personal health information (PHI), individual-level BCM-HM genetic and metabolomic data are available on request from the corresponding author (JMS). Computational code and pipelines used for data analysis are available in GitHub (https://github.com/ruthbpaula/PD_multiomics/).

## Results

### PD risk variant at *SPTSSB* associates with sphingolipid and fatty acid changes in blood

Common or rare variants in several genes, including *GBA1*, strongly implicate sphingolipid and ceramide metabolism in PD risk and pathogenesis (**Figure 1A**). We generated comprehensive metabolic profiles from blood (plasma) in 149 PD cases and 150 neurologically-healthy controls. PD cases had a mean age of 65.3 years (range 34-85), were 36.9% female, and had a mean disease duration of 8 years (**Supplemental Table 1**). Overall, we identified 62 sphingolipid and ceramide species, of which 26 (42%) showed suggestive PD-associated perturbations in blood (*p* < 0.05) (**Figure 1B**, **Supplemental Figure 3**, and **Supplemental Data 1**). Interestingly, some ceramide precursors were down-regulated in PD, whereas other complex sphingolipids were up-regulated (15 out of 18 total metabolites, including glucosylceramide and lactosylceramide). Our results are consistent with prior studies showing sphingolipid perturbations in PD^20^.

To differentiate proximal, causal changes from downstream metabolic perturbations resulting from PD, we restricted our analyses to controls without PD and further stratified the sample based on the presence of PD risk variants in genes that regulate sphingolipid/ceramide synthesis or breakdown (**Figure 1A**). *GBA1* variants are uncommon in our sample (only 9 PD cases and 6 controls), limiting statistical power to interrogate for differential metabolic signatures. Similarly, rare loss-of-functions variants in *SMPD1* or *ARSA*, which are also established to increase PD risk^7–9^, were not present in our dataset. We next considered common risk variants identified from PD genome-wide association studies^5,42^, including *rs28628748* (freq. = 0.37) in *SCARB2*, encoding a chaperone required for GBA1 transport to the lysosome, and *rs979812* (freq. = 0.44) at the *GALC* locus, which is linked to the lysosomal storage disorder, Krabbe disease. However, neither of these variants were associated with sphingolipid perturbations in our data.

Though not as well studied as *GBA1* and other lysosomal genes, a genome-wide significant association (*rs1450522^G^*, freq. = 0.31, *p* = 5 x 10^-10^) has also been reported^5^ at the *serine palmitoyltransferase small subunit B* (*SPTSSB*) locus, encoding a regulatory subunit of SPT, which participates in the initial, rate-limiting reaction for *de novo* sphingolipid synthesis. The SPT complex catalyzes the condensation of serine with a fatty acid derivative, usually palmitoyl-CoA, to generate 3-ketosphinganine (also known as 3-ketodihydrosphingosine)^52^. Interestingly, besides the association of common *SPTSSB* variants with PD risk, rare gain-of-function mutations in genes encoding other SPT subunits have been identified as causes of hereditary sensory and autonomic neuropathy type 1 (*SPTLC1* and *SPTLC2*), hereditary spastic paraplegia (*SPTSSA*), and amyotrophic lateral sclerosis / frontotemporal dementia (*SPTLC1* and *SPTLC2*)^14,53–56^. Among neurologically healthy controls in our local cohort, we identified 60 heterozygous and 9 homozygous carriers of the *SPTSSB^rs1450522-G^* PD risk allele, along with 79 non-carriers. Strikingly, we discovered that the *SPTSSB* variant was in fact associated with changes in of 8 out of 62 assayed sphingolipids and ceramides in our dataset (*p* < 0.05), including 4 out of the 26 sphingolipids altered in PD (above) (**Figure 1C**, **Supplemental Figure 5**, **Supplemental Table 3**). Notably, elevated plasma levels for glucosylceramide, lactosylceramide and multiple sphingomyelin species were observed among those with the *SPTSSB^rs14505^*^22^*^-G^* PD risk allele. For selected sphingomyelin and glucosyl ceramide species, the association appeared to be driven by the *SPTSSB^rs14505^*^22^*^-G^* homozygote class. Among PD cases, the association was further enhanced, such that *SPTSSB^rs1450522-G^* was associated with perturbations in 16 out of 62 sphingolipids (**Supplemental Table 3** and **Supplemental Figure 4**). Our results are potentially consistent with a metabolic quantitative trait locus (mQTL), linking *SPTSSB* genetic variation with increased sphingolipid production.

To further confirm the association between the *SPTSSB* variant and elevated sphingolipids, we leveraged available plasma lipidomics or metabolomics along with genotyping data from other published studies. We first considered data from an Australian cohort (*n* = 4,492) without known neurological disease ^51^. Strikingly, among 629 lipid species interrogated, the top-ranked associations with the *SPTSSB^rs14505^*^22^*^-G^* PD risk allele pinpoint increases in multiple ceramides [e.g., d20:1/24:0 (*p* = 0.005); d20:1/24:1 (*p* = 0.006)], dihydroceramide [d18:0/18:0 (*p* = 0.006)], as well as sphingomyelin species (**Figure 1D** and **Supplemental Data 3**). Consistent with increased *de novo* sphingolipid synthesis, we also discovered a suggestive association with elevated sphinganine levels (*p* = 0.025) in a large European cohort (n = 19,994 healthy individuals)^44^. Sphinganine is only two steps away from the reaction catalyzed by the SPT enzymatic complex (**Figure 1A**). Of note, only 23 sphingolipids and no ceramides were included in this European dataset. Lastly, to determine other candidate metabolic markers for *rs1450522* carriers, we interrogated all 301 lipids detected in this larger and more powerful cohort. Interestingly, these analyses revealed evidence for a potential broader impact of *SPTSSB^rs14505^*^22^*^-G^* on lipid metabolism, including reduced levels of the fatty acid, heptanoate (*p* = 6x10^-4^), among other species. Notably, fatty acids are important for sphingolipid synthesis, and the SPT substrate and fatty acid derivative, palmitoyl-CoA, is also a key building block and regulator of fatty acid synthesis^14,25^. We also found that the levels of selected sphingolipids and free fatty acids were correlated in both blood and brain tissue (**Supplemental Figures 10-11** and **Supplemental Data 1**).

#### PD risk variants colocalize with *SPTSSB* expression in brain and fatty acid perturbations in blood

The lead PD risk variant at *SPTSSB*, *rs1450522^G^*, is located within the 5’-untranslated region (UTR), suggesting a potential impact on mRNA processing and/or stability. Consistent with this, an *SPTSSB* expression quantitative trait locus (eQTL) was previously reported as part of systematic follow-up analyses from the genome-wide association study discovering this PD risk locus^5,57^. To further investigate the relation between *rs1450522* and *SPTSSB* mRNA expression we leveraged several complementary datasets pairing genotyping and RNA-sequencing (RNA-seq). In the Genotype-Tissue Expression Portal (GTEx)^58^ and BrainMeta^43^ datasets, we confirmed that rs1450522*^G^* is associated with increased *SPTSSB* expression in both bulk human brain and tibial nerve tissue (**Supplemental Figure 1A**). We also interrogated a single nucleus expression dataset from the Religious Orders Study and Rush Memory and Aging Project (ROSMAP)^49^ postmortem dorsolateral pre-frontal cortex tissue, highlighting a significant *SPTSSB* eQTL present in excitatory and inhibitory neurons, but not glia (**Supplemental Figure 1B-C**, **Supplemental Table 4**)^59^. The association was strongest in excitatory neurons, highlighting a potential dose-dependent relationship between the rs1450522*^G^* variant and *SPTSSB* mRNA levels (*p* = 8.1 x 10^-32^).

Together, our results show that rs1450522*^G^* is not only associated with PD risk, but also both increased *SPTSSB* expression and altered metabolism affecting multiple sphingolipids, fatty acids, and derivatives. To further examine the potential causal chain between *SPTSSB* genetic variation, gene expression, metabolism, and PD (**Figure 2A**), we next performed complementary analyses using colocalization followed by summary-data-based mendelian randomization (SMR) to examine for consistent, correlated and causal associations between genetic variation at the *SPTSSB* locus and the pertinent eQTL and mQTLs detected above (**Figure 2B**, **Table 1**, **Supplemental Table 5**, **Supplemental Data 3**). For optimal statistical power, we leveraged updated genome-wide association study summary statistics from the Global Parkinson’s Disease Genetics consortium (GP2)^42^, incorporating results from 64,047 PD cases and 1,754,191 controls. We primarily focused on the cell-type specific brain eQTL from excitatory neurons and the plasma heptanoate mQTL, comprising the most robust associations from our analyses of *SPTSSB^rs14505^*^22^*^-G^*.

**Figure 2.**
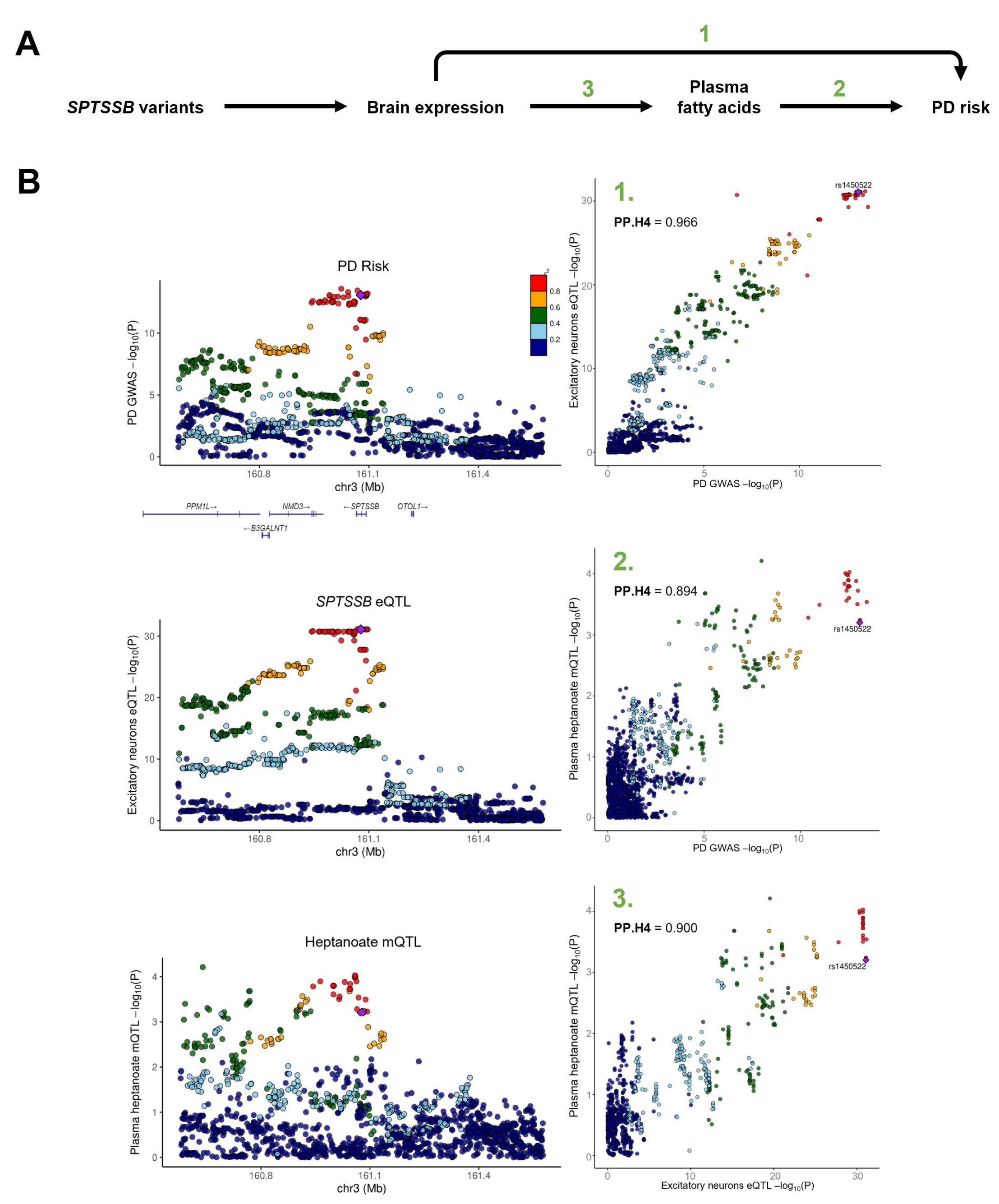
Parkinson’s disease (PD) risk variants at the *SPTSSB* locus co-localize with *SPTSSB* mRNA expression and fatty acid metabolism. (A) Hypothetical causal chain linking genetic variation at the *SPTSSB* gene with PD risk. (B) At left, locus plots (chromosome position versus p-value) show variant associations at the *SPTSSB* locus with PD risk (top, GP2 GWAS data), *SPTSSB* mRNA (middle, eQTL from ROSMAP excitatory neurons), or plasma heptanoate levels (bottom, mQTL from Surendran et al., 2022). At right, co-localization plots show causal relationships keyed to numbering in panel A. Significance testing based on posterior probability, PP.H4, denoting the likelihood that both traits share a common causal variant (PP.H4 > 0.5 was considered significant). Color key denotes linkage disequilibrium (LD) of each variant with *rs1450522*. See also Supplementary Data File 3.

**Table 1.** Summary-based-Mendelian randomization at the *SPTSSB* locus.

| Exposure | Outcome | Effect | SE | $P_{single}$ | $P_{multi}$ | $P_{HEIDI}$ |
| --- | --- | --- | --- | --- | --- | --- |
| *SPTSSB* eQTL (excitatory neurons) | PD | 0.24 | 0.04 | $1.94 \times 10^{-10}$ | $1.94 \times 10^{-10}$ | 0.59 |
| heptanoate mQTL (plasma) | PD | -1.82 | 0.53 | $5.71 \times 10^{-4}$ | $5.71 \times 10^{-4}$ | 0.52 |
| *SPTSSB* eQTL (excitatory neurons) | heptanoate mQTL (plasma) | -0.12 | 0.03 | $5.32 \times 10^{-4}$ | $3.54 \times 10^{-3}$ | 0.30 |

Summary-based Mendelian randomization (SMR) significance based on single and multi-SNP (multi) $p < 0.05$ , and HEIDI $p > 0.05$ to confirm the absence of pleiotropy.

We discovered significant colocalization between the association signal for PD genetic risk at the *SPTSSB* locus with either *SPTSSB* mRNA expression in excitatory neurons and plasma heptanoate levels [posterior probability (PP.H4) of 97% and 89% respectively], and similar results are seen for a direct comparison between *SPTSSB* eQTL and plasma heptanoate (PP.H4 = 90%) (**Figure 2** and **Supplemental Figure 6**). Consistent results were also independently obtained using SMR (**Table 1** and **Supplemental Data 3**). Overall, our results provide strong support for the hypothesis that the 3 traits (PD, excitatory neuron *SPTSSB* eQTL, heptanoate mQTL) share a common causal variant, consistent with a molecular cascade from common genetic variation at *SPTSSB* to increased mRNA expression, altered metabolism, reduced plasma heptanoate levels, and PD susceptibility. In addition, we obtained consistent, albeit somewhat attenuated results when using dorsolateral prefrontal cortex bulk tissue instead of the single-nucleus expression profiles from excitatory neurons (**Table 1**, **Supplemental Figure 7**, **Supplemental Data 3**). However, it was not possible to robustly model the causal pathway within a single tissue compartment (i.e., brain or blood) (**Supplemental Figure 1A**). Interestingly, in data from one recent published analysis^26^, we did find a modest association (*p* = 0.035) between *SPTSSB^rs1450522-G^*and increased glucosylceramide in cerebrospinal fluid. Lastly, we also considered other candidate genes at the susceptibility locus defined by *rs1450522* (**Supplemental Data 3**). Besides *SPTSSB* and consistent with published results^5,57,60^, we also confirmed an eQTL for the adjacent gene, *NMD3*, which was present in blood and bulk cortical brain tissue but not excitatory neurons. NMD3 regulates RNA processing and is not known to have a role in lipid metabolism. Other nearby genes, including *PPM1L*, *B3GALNT1*, and *OTOL1*, did not show any evidence for an eQTL.

#### PD is characterized by fatty acid and acylcarnitine perturbations in blood

The preceding analyses highlight how sphingolipids and other fatty acid species might serve as both PD biomarkers and causal mediators, linking genetic risk variants to disease manifestations. In order to systematically define other promising PD signatures, we next conducted a metabolome-wide analysis, interrogating 1,171 metabolites from blood plasma (**Supplemental Figure 2**) among our PD case/control cohort. In this exploratory analysis, we identified 327 differential metabolites meeting a suggestive significance threshold (*p* < 0.05), including 97 increased and 230 decreased species (**Figure 3A-B**, **Table 2**, **Supplemental Data 1**). The most extreme, PD-associated metabolic perturbations in blood, are well-known breakdown products of levodopa^61^, including 3-methoxytyrosine, vanillactate, 3-methoxytyramine sulfate, and other tyrosine derivatives (**Figure 3A**). Alterations in plasma lipids were the most frequent among all PD-associated metabolic changes (86 out of all 327 differential metabolites), possibly due to the disproportionate representation of this class in our assay panel (**Figure 3C**, **Supplemental Figure 2)**. As expected, the sphingolipid pathway was enriched for PD-associated changes, particularly among sphingomyelins, with 7 increased and 2 decreased metabolites (**Figure 3D**). Behenoyl-dihydrosphingomyelin was one of the top-ranked downregulated species in the PD metabolome, with a mean 0.71-fold decrease in PD plasma (*p* = 2.8 x 10^-11^).

**Figure 3.**
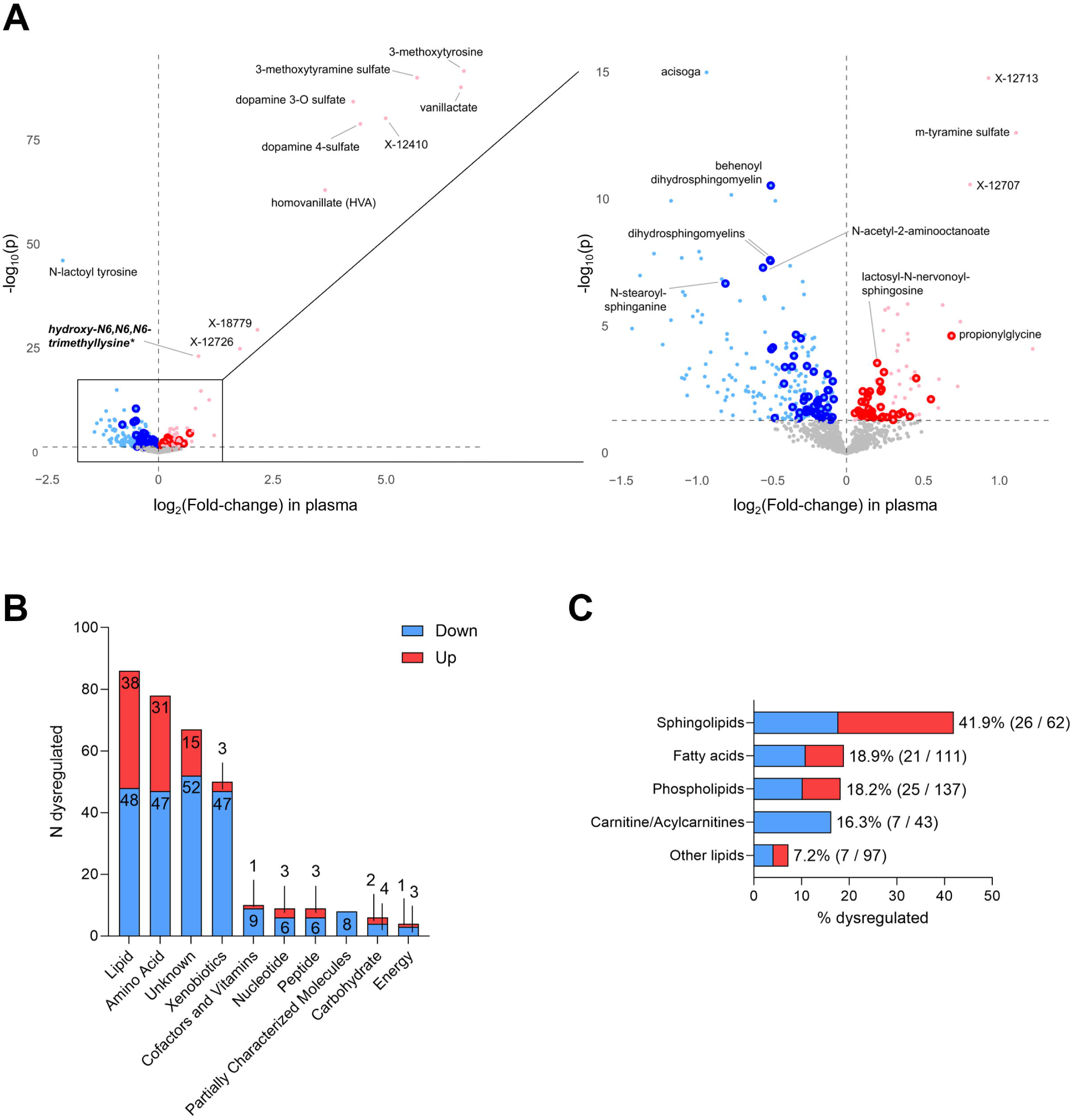
Parkinson’s disease (PD) is characterized by lipid metabolic perturbations in blood. Differential metabolism was assessed in plasma from 149 PD cases and 150 controls without PD, using the likelihood ratio test and adjusting for age and sex. (A) Volcano plots show up- (red) or down-regulated (blue) meeting the suggestive significance threshold (unadjusted *p <* 0.05 dashed horizontal line). At left, the full plot is notable for extreme perturbations in levodopa metabolites. At right, these outlier values are excluded to magnify the boxed region. Lipid species are denoted with larger, darker dots. See also Table 2. (B) Plots showing the total with number of dysregulated species in PD, broken down by metabolite class. Up- and down-regulated, denoted by red and blue color, respectively. (D) Plots showing the proportion (%) of dysregulated species within each lipid class, with up- (red) and down- (blue) regulated changes indicated. The sphingolipid and other lipid classes were significantly enriched for dysregulated metabolites, based on a Fisher test *p* < 0.05.

**Table 2.**
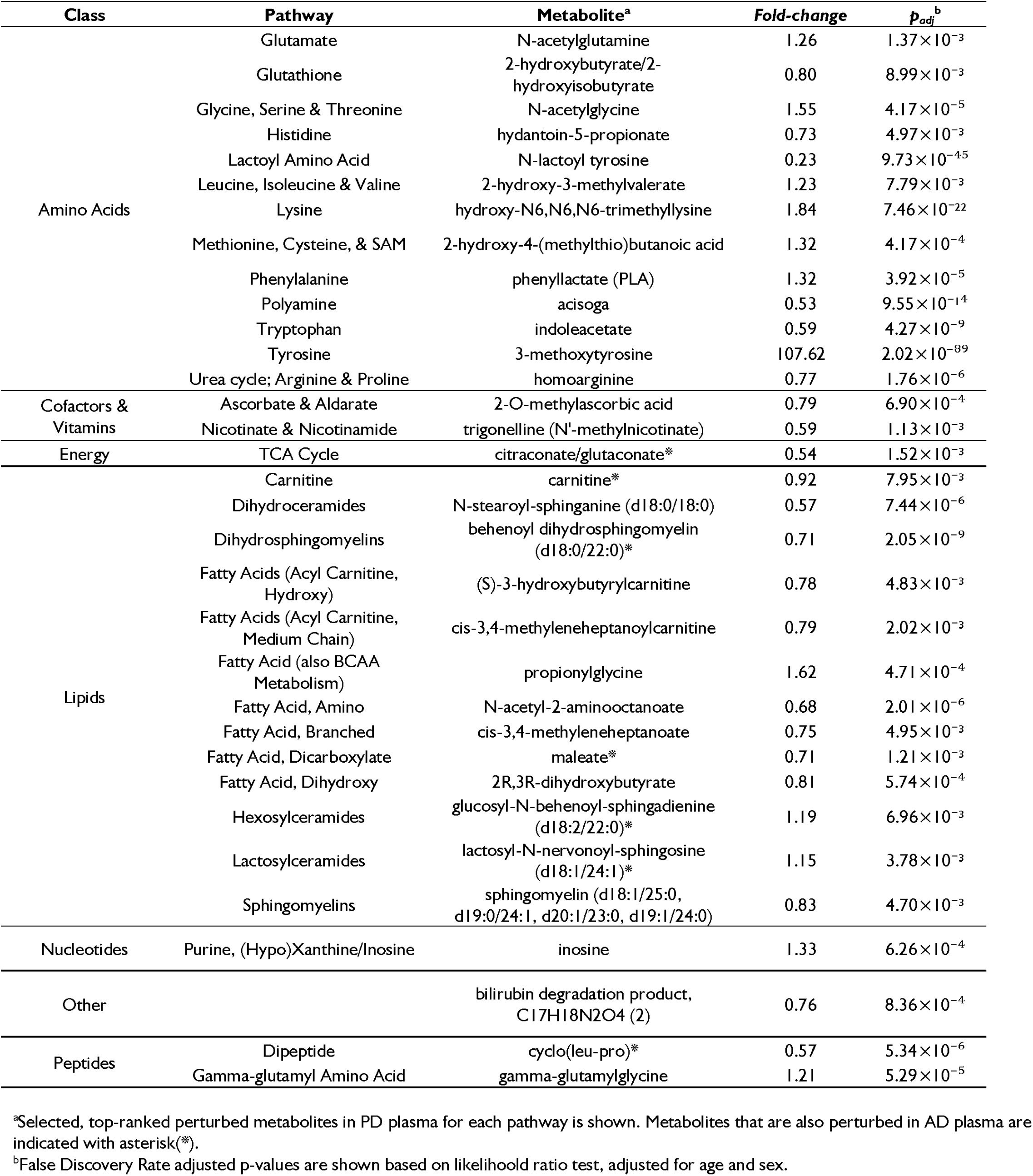
Representative perturbed metabolites in PD plasma.

We also detected PD-associated plasma perturbations in several phospholipids (11 increased / 14 decreased), free fatty acids (9 increased / 12 decreased), and acylcarnitines (7 decreased). Although heptanoate was not detected in our dataset, the closely related cis-3,4-methyleneheptanoate was decreased in PD and was the most significantly perturbed branched fatty acid (fold-change = 0.75; *p* = 4x10^-4^). Acylcarnitines, which are comprised of fatty acids linked to a carnitine moiety, have important functions in mitochondrial beta-oxidation and have previously been reported to be reduced in blood from individuals with PD^16,61,62^. For example, we documented PD-associated reductions in cis-3,4-methyleneheptanoylcarnitine (fold-change = 0.79, *p* = 0.0001), (S)-3-hydroxybutyrylcarnitine (fold-change = 0.78, *p* = 0.0004), and carnitine itself (fold-change = 0.92, *p* = 0.0007). Interestingly, the top-ranked, up-regulated metabolite in PD is an amino acid derivative, hydroxy-N6,N6,N6-trimethyllysine, linked to carnitine synthesis^63^.

To determine whether metabolic perturbations are specific for PD, we also analyzed blood from an additional 99 individuals with Alzheimer’s disease (AD) (**Supplemental Figure 9** and **Supplemental Data 1**). All samples were analyzed together with the PD cases and compared with the same control group. Overall, we identified significant overlap between the differential blood metabolic signatures of AD and PD (*p* = 2.2x10^-11^) (**Supplemental Figure 9A**). For example, sphingolipid perturbations were also detected in AD, with 18 overlapping and concordant metabolites between AD and PD, including 5 down- and 13 up-regulated species (**Supplemental Data 1**). Further, 3 out of the 7 acylcarnitines down-regulated in PD were consistently and concordantly perturbed in AD. However, whereas PD showed a uniformly decreased acylcarnitine signature, 8 additional acylcarnitine species were increased selectively in AD.

Besides lipids, our metabolome-wide analyses also identified PD-specific blood metabolite changes representative of dysregulation in the tricarboxylic acid and urea cycles, as well as altered metabolism of amino acids, polyamines, and nicotinamide metabolism (**Table 2**; **Supplemental Data 1**). On the other hand, potential AD-specific changes included perturbations affecting long chain fatty acids, lysophospholipids, secondary bile acids, and sterols, consistent with findings from other published work^64–66^.

#### A brain acylcarnitine metabolic signature also accompanies preclinical alpha-synuclein pathology

We next investigated whether PD metabolic signatures from blood can also be detected in the brain. The diagnosis of PD is preceded by a decades-long, clinically silent phase in which alpha-synuclein brain neuropathology develops in association with absent or minimal motor impairment^1^. Therefore, it will be essential to define biomarkers indicating the development and progression of preclinical PD pathophysiology—these markers may also enhance understanding of the causal chain linking the earliest disease triggers to its clinical manifestation. To characterize metabolic changes associated with alpha-synuclein Lewy body pathology, we again leveraged data from ROSMAP^29^, a prospective study of brain aging that enrolls older adults for longitudinal clinical evaluations with brain autopsy and comprehensive neuropathologic studies following death.

Our analysis included a total of 490 brain autopsies (90.5 y mean age of death, 70.4% female), including 129 cases with Lewy body (LB) pathology and 361 controls without LB pathology (**Supplemental Table 2**). Metabolome profiles from postmortem dorsolateral prefrontal cortex tissue were available for analysis (*n* = 685 total metabolites), based on the same comprehensive assay as in our PD clinical cohort with plasma metabolomics. We interrogated for differential metabolite perturbations associated with LB pathology following adjustment for age, sex, and postmortem interval. Despite the increased sample size, we detected ∼3-fold fewer metabolite perturbations in postmortem brain versus blood, when considering LB pathology versus PD clinical diagnosis, respectively. We speculate that metabolic signatures may be rapidly attenuated and potentially obscured due to postmortem artifact, as we noted a strong association between many metabolites and postmortem interval (**Supplemental Data 1**). Nevertheless, 68 total metabolites were differentially detected in brains with LB pathology, meeting our suggestive significance threshold (*p* < 0.05) (**Figure 4A**). Of those, 15 out of 68 (22%) overlapped with perturbations in blood from our PD case-control analysis, of which two-thirds showed concordant changes (**Supplemental Data 1**).

**Figure 4.**
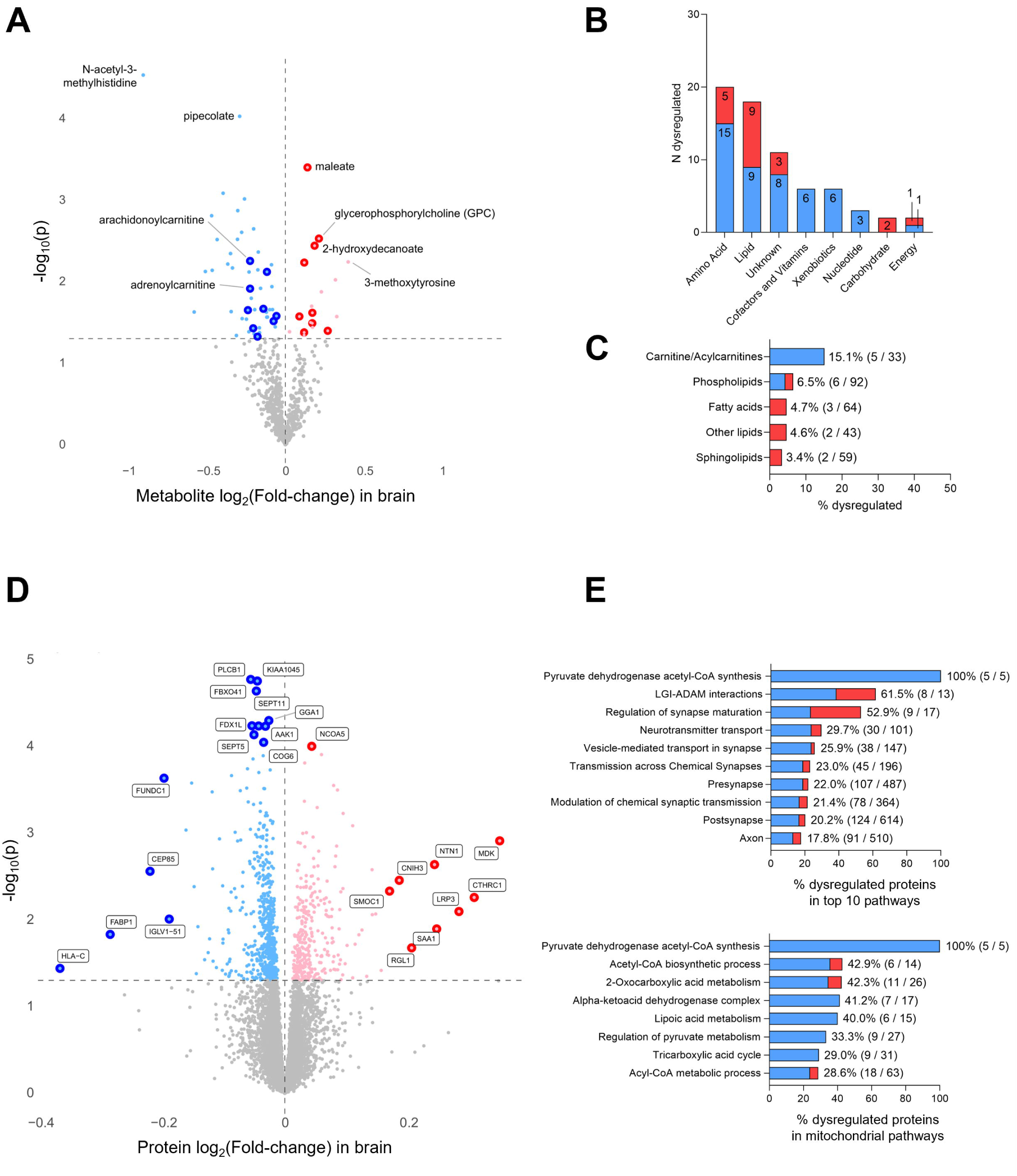
Alpha-synuclein Lewy body (LB) pathology is characterized by an acylcarnitine metabolic signature and differential expression of mitochondrial proteins. Differential metabolism and protein expression were interrogated in ROSMAP postmortem brain tissue (dorsolateral prefrontal cortex) data including 129 cases with LB pathology and 361 controls without LB pathology, using the likelihood ratio test and adjusting for age and sex. Volcano plots show up- (red) or down-regulated (blue) metabolites (A) or proteins (D) meeting the suggestive significance threshold (unadjusted *p < 0.05* dashed horizontal line). Selected metabolites or proteins are labeled. See also Supplemental Data 2. Plots showing the number (B) and proportion (C) of dysregulated species associated with LB pathology, broken down by metabolite class. Up- and down-regulated, denoted by red and blue color, respectively. The carnitine/acylcarnitine class was significantly enriched for dysregulated metabolites, based on a Fisher test *p* < 0.05. (E) Plot showing results of pathway enrichment analysis from differential protein expression in LB pathology, including top-ranked pathways (top) and mitochondrial pathways (bottom). The proportion (%) of differential expressed proteins from pathway is noted, with up- (red) and down- (blue) regulated changes indicated. (%). All displayed pathways were significantly enriched, based on Fisher test (false discovery rate adjusted *p* < 0.05).

After amino acids, lipid perturbations accounted for the second largest fraction of changes detected in LB positive cases (18 out of 68 metabolites, 37%) (**Figure 4B**). However, in the brain, sphingolipids were no longer the dominant signature, with only two sphingolipid metabolites altered [glycosyl-N-tricosanoyl-sphingadienine (d18:2/23:0) and lactosyl-N-stearoyl-sphingosine (d18:1/18:0)]. Instead, carnitine and acylcarnitine metabolites were the most enriched species showing changes in cases with alpha-synuclein pathology (**Figure 4C**). A total of 5 acylcarnitines were decreased in brains with LB pathology, including arachidonoyl-carnitine (fold-change = 0.86, *p* = 0.006) and adrenoylcarnitine (fold-change = 0.86, *p* = 0.01). When considering all acylcarnitines detected by the assay, regardless of significance in the differential analysis, we found evidence for systematic down-regulation among this lipid class in brains, with 28 out of 33 detected species reduced, making this finding unlikely to be a chance finding (*p* = 1 x 10^-5^).

Importantly, only 20 of the 129 cases with alpha-synuclein LB pathology included in our analysis carried a PD diagnosis, consistent with a predominantly preclinical or prodromal cohort^67^. In a secondary analysis, we excluded the small number of LB pathology cases with a clinical diagnosis of PD (**Supplemental Data 1**). As expected, the major levodopa breakdown product^68^, 3-methoxytyrosine, which was present in the primary analysis, was no longer detected in this secondary analysis. Interestingly however, the acylcarnitine signature was preserved in this analysis, strongly suggesting decreases in this class of lipids may accompany PD pathology prior to the manifestation or recognition of the full PD clinical syndrome.

#### Alpha-synuclein pathology is associated with a brain proteomic signature suggestive of mitochondrial dysfunction

Our results, along with other published work^13,16–23^, strongly suggest that PD pathology and clinical manifestations are accompanied by altered lipid metabolism in brain and blood, and that certain sphingolipid and fatty acid signatures may mediate the impact of risk genes like *SPTSSB*. To further elaborate the responsible mechanisms for lipid perturbations in PD, we examined differential protein expression in the setting of alpha-synuclein pathology, leveraging available mass-spectrometry proteomics from the same brain autopsy cohort used for metabolomics (93 LB cases and 293 controls) (**Figure 4D-E** and **Supplemental Data 2**).

Overall, we detected perturbations in 896 proteins after adjustment for age at death, sex, and postmortem interval (*p* < 0.05), including 314 up- and 582 down-regulated proteins. Interestingly, the top LB-associated, upregulated protein was midkine (MDK; fold-change = 1.27; *p =* 0.001), a secreted growth and immunomodulatory factor, that has also been implicated from AD proteomics^69,70^. Based on overrepresentation analyses, LB pathology was characterized by a protein expression signature enriched for biological pathways implicated in synaptic structure/function and vesicle trafficking (**Figure 4E**, **Supplemental Data 2**), consistent with recent published proteomic studies of postmortem brain tissue from LB dementia^71^. Interestingly, we also noted significant enrichment for mitochondrial processes, bioenergetics, and fatty acid metabolism. Indeed, a preponderance of mitochondrial proteins were decreased in brains with LB pathology [e.g., CPT1A; fold-change = 0.98; *p* = 0.01] (**Supplemental Table 6**). Since acylcarnitines have a critical role in the transport of fatty acids into mitochondria for beta-oxidation^25^, our complementary metabolomic and proteomic analyses may identify convergent signatures related to mitochondrial dysfunction in PD.

## Discussion

A major goal for our field is to understand the molecular cascade from genetic variants to PD pathogenesis, since these pathways may identify promising biomarkers and targets for disease modifying therapies. *GBA1* and several other risk genes strongly implicate perturbations in sphingolipid/ceramide metabolism in PD pathogenesis^5–9^. We have integrated genomic, metabolic, transcriptomic, and proteomic data, including from blood and postmortem brain tissue, to probe mechanisms of PD risk and pathogenesis.

Building on recent genome-wide association studies, we discovered evidence for a causal chain at the *SPTSSB* susceptibility locus, linking genetic variants, increased mRNA expression, altered sphingolipid and fatty acids levels, and PD risk. *SPTSSB*, encoding a regulatory subunit for the SPT enzyme which catalyzes the initial step in *de novo* sphingolipid biosynthesis, has not been previously studied in PD. SPT condenses a fatty acyl-CoA with serine to create 3-ketosphinganine, which is subsequently modified to form the ceramide backbone for sphingolipids (**Figure 1A**). The trimeric SPT holoenzyme is comprised of 2 large subunits, SPTLC1 and either SPTLC2 or SPTLC3, along with the small regulatory subunit, either SPTSSA or SPTSSB. SPTSSB modifies SPT substrate specificity, favoring the generation of longer chain ceramides^52,72^. Whereas palmitoyl-CoA produces an 18-carbon chain base, which is the most abundant form, alternate substrates, such as stearoyl-CoA or myristoyl-CoA, can lead to longer (C20) or shorter (C16) backbones, respectively, contributing to a diversity of sphingolipid species with varying ceramide backbone lengths^73^. In a naturally occurring gain-of-function mouse mutant, *Stellar*, a single amino acid change in *SPTSSB* (His56Leu) increases the production of longer, C20 backbone sphingolipids, leading to intracellular membrane-like inclusions and neurodegeneration^74^. While the causal variant for PD risk remains to be established, the lead variant from published GWAS, *rs1450522*, falls in the *SPTSSB* 5’-UTR, and could potentially impact mRNA turnover. We hypothesize that increased *SPTSSB* expression promotes PD pathogenesis by either increasing overall sphingolipid production and/or favoring the generation of ceramide species with longer base chain lengths, either of which may have a deleterious impact on the central nervous system.

*SPTSSB* is expressed in brain, and we confirmed strong colocalization for PD risk variants with *SPTSSB* mRNA expression in excitatory neurons^5,57^. Whereas *SPTSSB^rs14505^*^22^ showed suggestive associations with increases in several blood ceramide and sphingolipid species, we did not detect similar associations with sphingolipids detected in postmortem brain tissue, nor was this variant related to brain fatty acids or other lipid species. It is possible that larger sample sizes will permit more powerful and direct analyses of brain sphingolipid metabolism; although, the rapid development of postmortem change may obscure metabolic signatures. It is also possible that some of the intermediate species in sphingolipid biosynthesis are short-lived, making them difficult to detect in brain or elsewhere. For example, 3-ketosphinganine the product of SPT, is rapidly transformed to sphinganine *in vivo*^75^. Since *SPTSSB* is poorly expressed in peripheral blood cells^76,77^, the sphingolipids altered in individuals with the *SPTSSB^rs1450522^* variant may represent spillover from brain sphingolipid pools, or alternatively, could derive from another unknown peripheral tissue source. Common genetic variants at *arylsulfatase A* (*ARSA*) show associations with both PD risk and reduced levels of O-sulfo-L-tyrosine in cerebrospinal fluid^26^, and we confirmed that this mQTL is similarly preserved in plasma (**Supplemental Figure 8**). We also documented significant colocalization between *SPTSSB* locus variants with PD risk and decreased plasma levels of the fatty acid heptanoate. As discussed further below, although sphingolipid and fatty acid metabolism have multiple plausible connections, additional work will be required to establish whether and how increased *SPTSSB* expression may secondarily trigger depletion of heptanoate or other fatty acids. Notably, the SPT substrate, palmitoyl-CoA, is a key intermediate and modulator for both fatty acid synthesis and breakdown^25^. Fatty acids are also consumed during the maturation of complex sphingolipids, to generate acyl side chains. Therefore, it is possible that increased SPT activity might indirectly lead to reduced fatty acids; although, other more complex feedback interactions are also possible.

In our metabolome-wide analysis, we discovered evidence for PD associated dysregulation in fatty acid metabolism, with reductions in multiple acylcarnitines observed in both blood and brain. Acylcarnitines mediate the transfer of fatty acids into mitochondria for breakdown via beta-oxidation^25^. Consistent with this, we found that PD pathology-associated differential protein signatures were significantly enriched for markers of mitochondrial dysfunction, including altered fatty acid metabolism. Based on several independent lines of evidence, mitochondria have long featured centrally in models for PD pathogenesis^1,78^.

Autosomal recessive, juvenile onset PD has been linked to several genes (e.g., *PARKIN/PARK2*) with roles in mitochondrial quality control^24^. Separately, toxins like rotenone and misfolded alpha-synuclein can similarly target the mitochondrial electron transport chain, promoting oxidative injury as a potential driver for neurodegeneration^79,80^. The convergent metabolic and proteomic signatures of PD and alpha-synuclein pathology are highly suggestive of altered bioenergetics. Our results also implicate perturbations in the TCA cycle (**Supplemental Table 6**), which is not only intimately related to mitochondrial respiration, but also involves key intermediates for both fatty acid and sphingolipid metabolism. The reciprocal interconnection among these metabolic networks is highlighted by experimental manipulations limiting serine bioavailability, which not only profoundly impact sphingolipid/ceramide synthesis (serine is a key substrate for SPT), but also trigger reduced acylcarnitine levels and impaired mitochondrial structure / function, including fatty acid oxidation^81^.

Our results add to prior studies highlighting lipid signatures, including sphingolipids^19–21^, fatty acids^16,22^, and fatty acid derivatives like acylcarnitines^61,62^ as promising blood biomarkers for PD. Fewer studies by comparison have also considered postmortem brain tissue, and most of these have been limited by small sample sizes. The autopsy cohort employed here, in which most cases with alpha-synuclein LB pathology are representative of potential early, preclinical PD or Lewy body dementia^67^, is particularly attractive for biomarker discovery. Another strength of our approach comes from the integration of metabolic and genetic data to pinpoint causal pathways. Abundant evidence from human genetics supports a causal pathway from loss-of-function in genes related to ceramide metabolism to PD risk; however, numerous studies also strongly suggest that alpha-synuclein and PD pathophysiology may reciprocally disrupt endosome-lysosomal function, amplifying sphingolipid dysmetabolism^1,12^. If correct, this bi-directional feedback loop could make it impossible to determine whether metabolic perturbations stem from genetic drivers or rather represent downstream effects of alpha-synuclein pathology. To circumvent this potential confounding, we examined *SPTSSB* variants in neurologically-healthy controls, pinpointing sphingolipid perturbations in individuals at risk for PD but without known pathology. In our complementary analyses of PD cases, *SPTSSB^rs14505^*^22^ was associated with an increased number of distinct but overlapping subset of sphingolipid metabolites (**Supplemental Table 3**). Notably, prior studies have found glucosylceramide and other sphingolipid changes in *GBA1-*PD cases, but most find that the changes overlap with those seen in idiopathic PD^18^. Thus, in future work, it may be important to interrogate metabolism in *GBA1* variant carriers without PD.

In sum, our results highlight promising lipid biomarkers for PD, and in the case of *SPTSSB*, we show how these signatures can aid in the dissection of causal pathways. It will be important to replicate our findings in larger samples, and to further examine sphingolipids, fatty acids, and acylcarnitines in relation to PD progression, heterogeneity, and therapeutic response. Confirmation of causal lipid signatures for PD risk may further open new therapeutic avenues. Pharmacologic manipulation of lipid metabolism is a well-established therapeutic strategy in cardiovascular disease prevention, making it highly feasible to consider for PD.

## Data Availability

ROSMAP data is available for download from the Synapse AD Knowledge Portal (https://adknowledgeportal.synapse.org), including bulk brain tissue metabolomics (syn26007830)28, bulk and single-cell RNA-sequencing (syn350572029, syn5336681849), proteomics (syn21449447)47, and whole genome sequencing (syn11707419)50. ROSMAP clinical, pathologic, and demographic data was also obtained from Synapse or requested directly from the Rush Alzheimers Disease Center. Data from the Surendran et al. metabolome-wide association analysis is accessible at (https://omicscience.org/apps/mgwas/mgwas.table.php); summary statistics for the SPTSSB locus were provided by Dr. Claudia Langenberg (University of Cambridge). We also downloaded summary statistics from the Cadby et al. lipidome-wide association analysis. Complete summary statistics from all genetic and metabolomic data from the BCM-HM cohorts are included with the supplemental data as part of this work. Due to privacy concerns and the possible inadvertent release of personal health information (PHI), individual-level BCM-HM genetic and metabolomic data are available on request from the corresponding author (JMS). Computational code and pipelines used for data analysis are available in GitHub (https://github.com/ruthbpaula/PD_multiomics/).

## Acknowledgements

We thank the patients and others from Baylor College of Medicine and Houston Methodist Hospital who generously participated in our research study. The results published here are in part based on data obtained from the AMP-AD Knowledge Portal (https://doi.org/10.7303/syn2580853). Additional data from the Religious Orders Study and Rush Memory and Aging Project were provided by the Rush Alzheimer’s Disease Center, Rush University Medical Center, Chicago. We also thank Dr. Claudia Langenberg from the University of Cambridge for sharing data from the Surendran et al. study, and the Global Parkinson’s Genetics Program (GP2) for providing GWAS summary statistics. GP2 is funded by the Aligning Science Across Parkinson’s (ASAP) initiative and implemented by The Michael J. Fox Foundation for Parkinson’s Research (MJFF). For a complete list of GP2 investigators, see https://gp2.org.

## Supplementary material

Supplementary Material includes 11 Figures, 6 Tables, and 3 Supplemental Data Files.

