## Supplemental Figures and Tables for "Mapping the causal chain from genetic risk variants to lipid dysmetabolism in Parkinson’s disease"

### Figure S1.

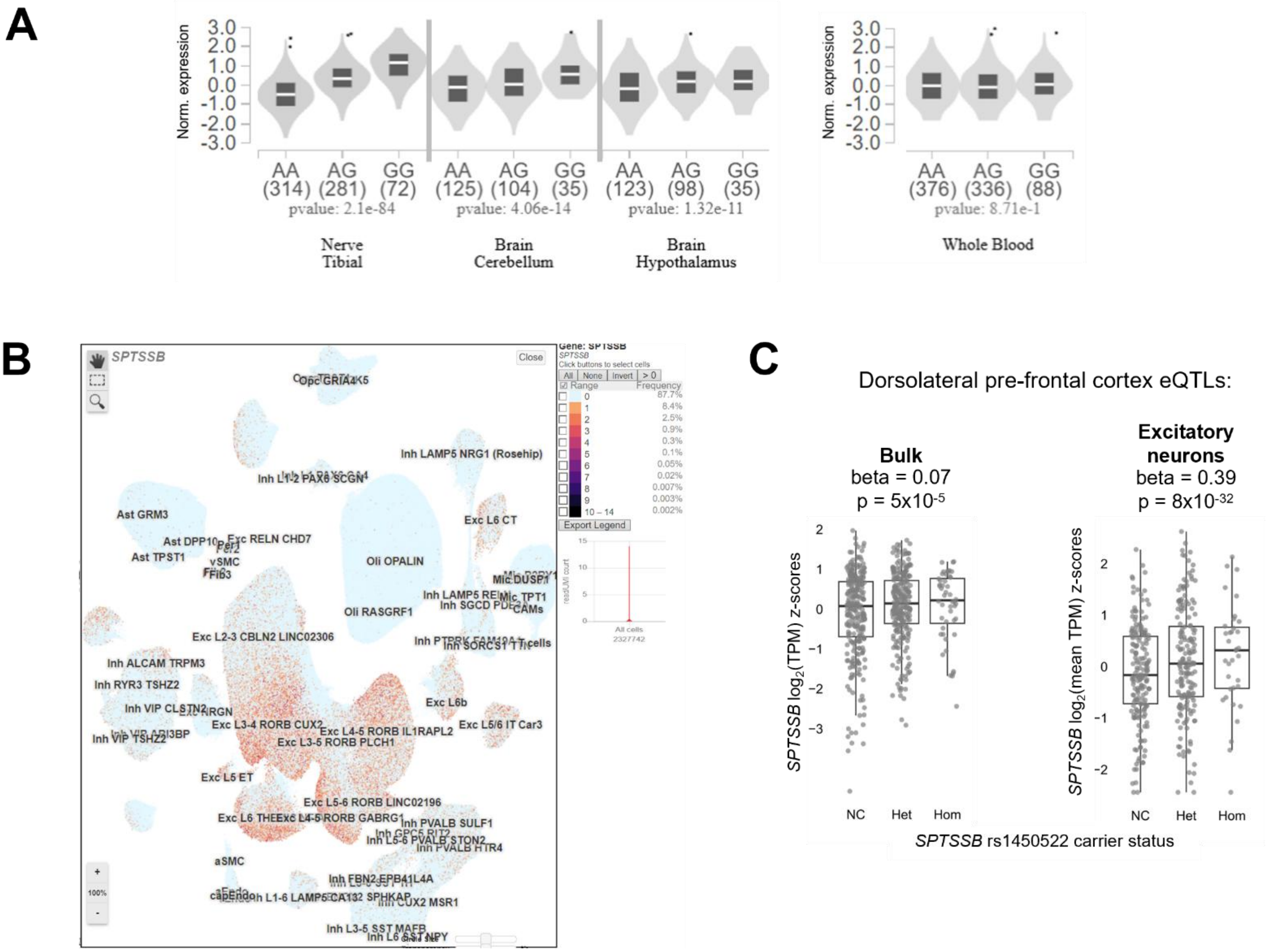


**Figure S1. Additional analysis of Parkinson's disease (PD) risk variants at the SPTSSB locus and *SPTSSB* mRNA expression.** (A) Based on GTEx data (https://gtexportal.org/), *SPTSSB* (rs1450522-G), which is associated with PD risk, is also associated with increased *SPTSSB* mRNA expression in tibial nerve, cerebellum, and hypothalmus, but not blood. (B) *SPTSSB* is strongly expressed in excitatory neurons, based on single-nucleus RNA sequencing data from ROSMAP dorsolateral prefrontal cortex samples (http://compbio2.mit.edu/ad_multiregion/). (C) Boxplots show significant expression quantitative trace loci (eQTL) for *SPTSSB* mRNA in bulk brain tissue from dorsolateral pre-frontal cortex as well as single-nucleus data from excitatory neurons. Data was extracted from the ROSMAP xQTLServer (https://mostafavilab.stat.ubc.ca/xqtl/) or summary statistics on Synapse (syn52335807).

**Figure S2.**

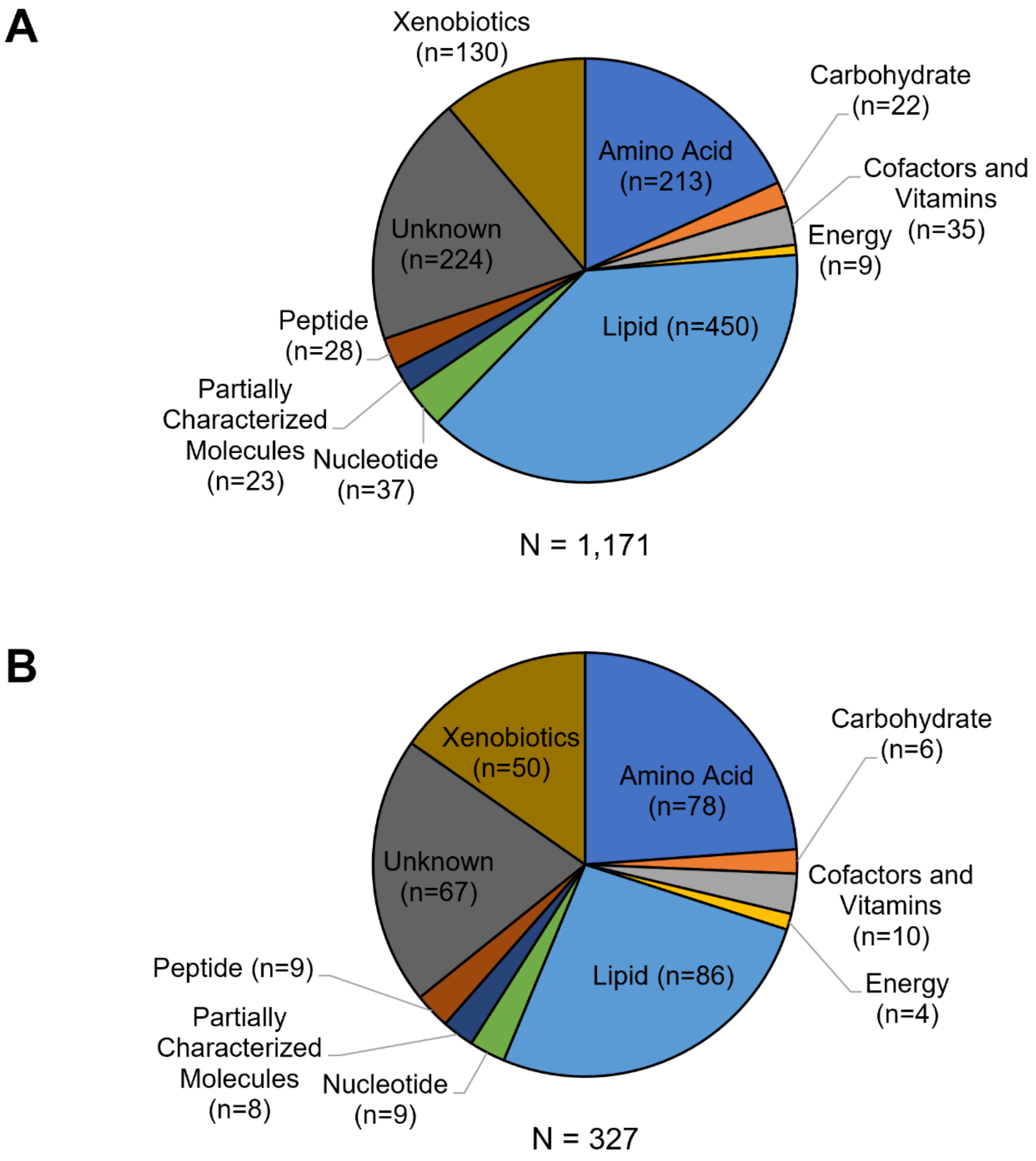


**Figure S2. Metabolic pathways interrogated in the Houston PD case/control cohort.** Pie charts displaying (A) all metabolites analyzed, and (B) metabolites perturbed in PD cases versus control (unadjusted $p < 0.05$).

### Figure S3.

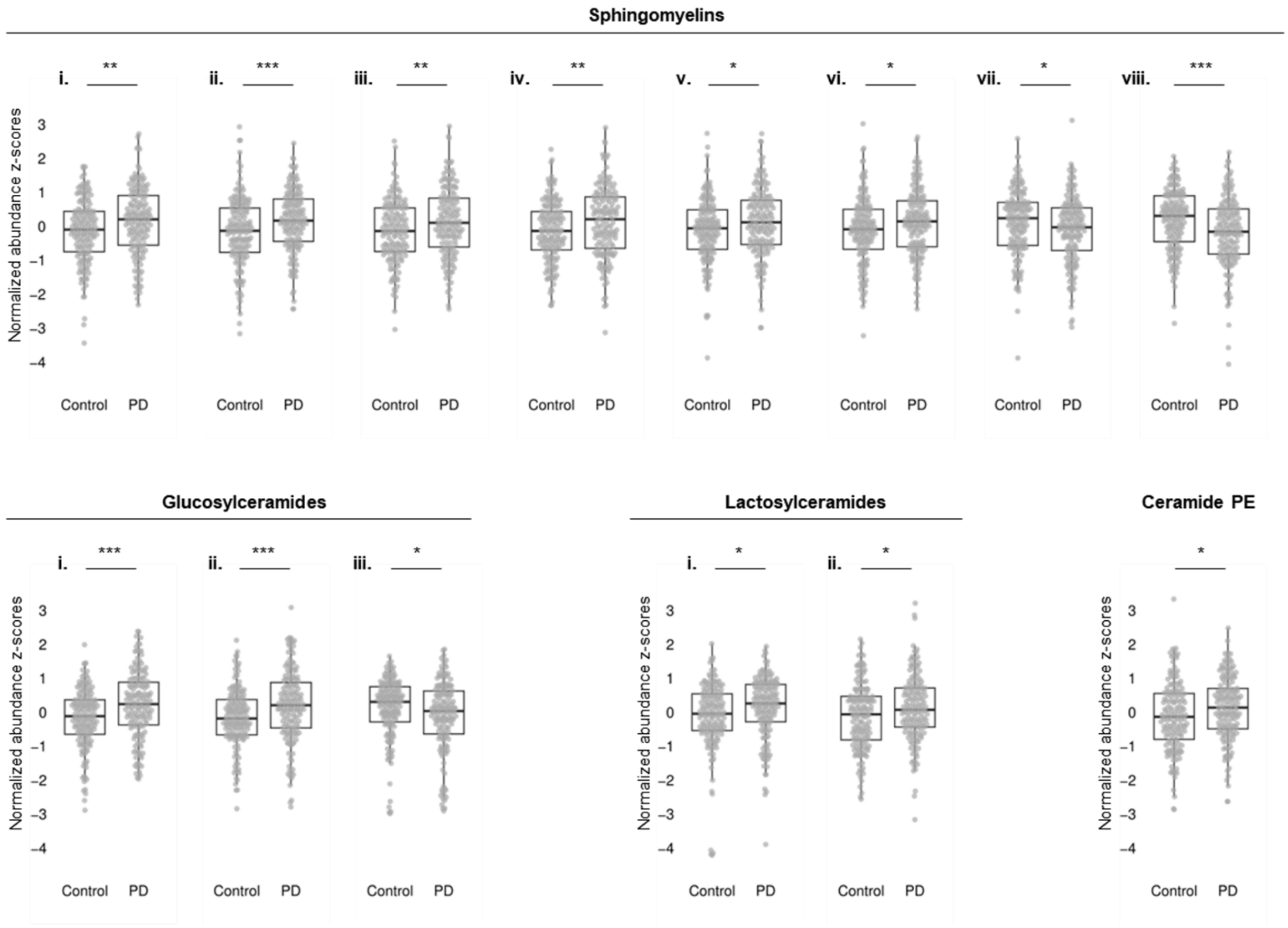


**Figure S3. Additional sphingolipids and ceramides perturbed in Parkinson's disease (PD).** Boxplots show additional sphingolipid perturbations from 149 PD cases versus 150 controls. Normalized residuals (z-scores) are plotted, following adjustment for age and sex. The whiskers denote the interquartile range between the first and third quartiles. Significance testing was based on likelihood-ratio tests (LRT). *, $p < 0.05$; **, $p < 0.01$; ***, $p < 0.005$. The following sphingomyelins are shown: (i) sphingomyelin (d18:2/24:2), (ii) hydroxypalmitoyl sphingomyelin (d18:1/16:0(OH)), (iii) sphingomyelin (d18:2/18:1), (iv) sphingomyelin (d18:1/20:1, d18:2/20:0), (v) sphingomyelin (d18:2/16:0, d18:1/16:1), (vi) palmitoyl sphingomyelin (d18:1/16:0), (vii) sphingomyelin (d18:1/21:0, d17:1/22:0, d16:1/23:0), (viii) sphingomyelin (d18:1/25:0, d19:0/24:1, d20:1/23:0, d19:1/24:0). The following glucosylceramides are shown: (i) glycosyl-N-behenoyl-sphingadienine (d18:2/22:0), (ii) glycosyl ceramide (d18:2/24:1, d18:1/24:2), (iii) glycosyl-N-(2-hydroxynervonoyl)-sphingosine (d18:1/24:1(2OH)). The following lactosylceramides are shown: (i) lactosyl-N-behenoyl-sphingosine (d18:1/22:0), (ii) lactosyl-N-palmitoyl-sphingosine (d18:1/16:0). Ceramide PE = palmitoyl-sphingosine-phosphoethanolamine (d18:1/16:0). See also Figure 1B for additional examples.

### Figure S4.

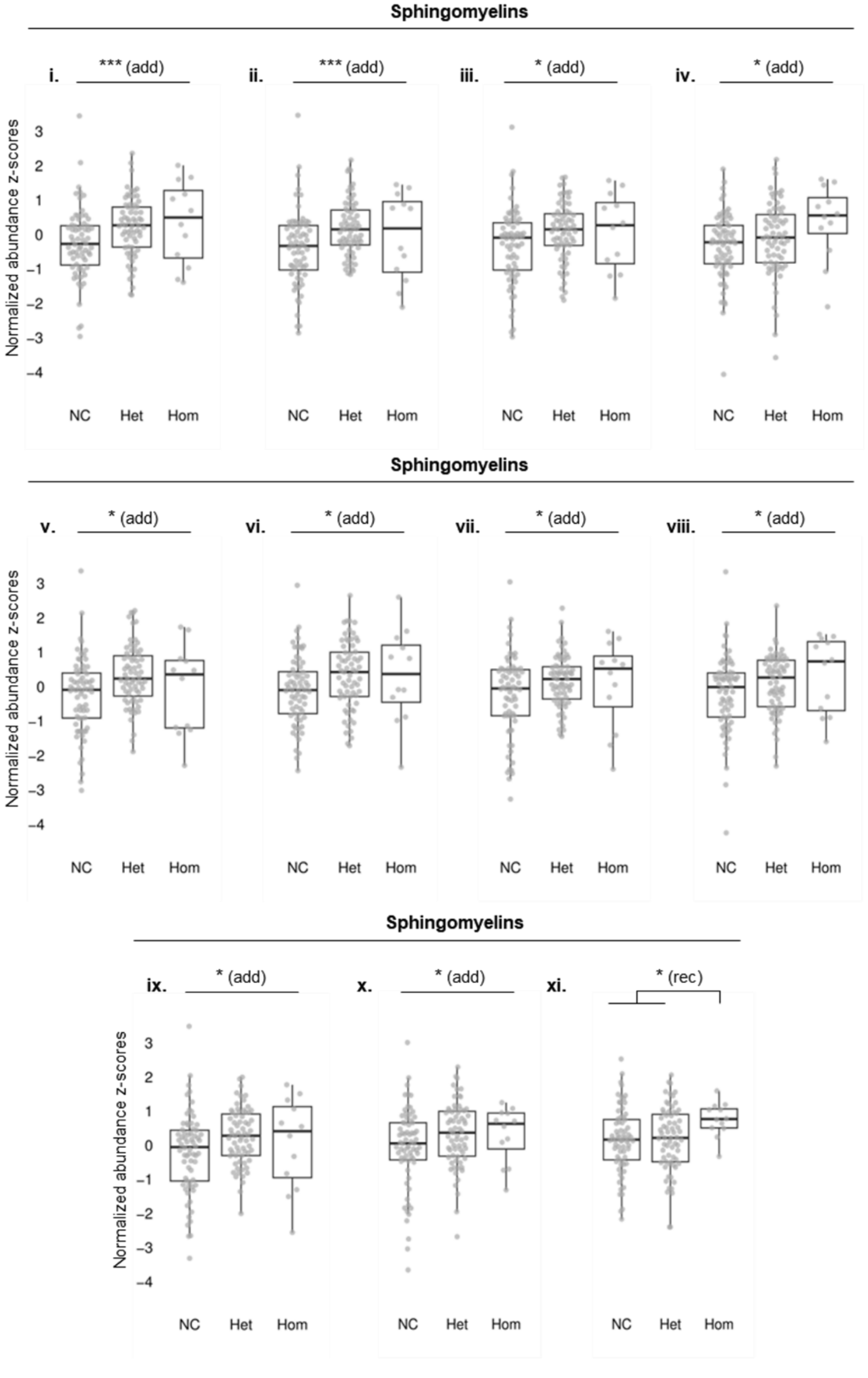


**Figure S4. Associations between *rs1450522* and plasma sphingolipids in PD cases.** The whiskers denote the interquartile range between the first and third quartiles. Normalized residuals (z-scores) are plotted, following adjustment for age and sex. Significance testing was based on logistic regression considering either an additive (add) or recessive (rec) inheritance model. *, $p < 0.05$; ***, $p < 0.005$. The following sphingomyelins are shown: (i) sphingomyelin (d18:1/17:0, d17:1/18:0, d19:1/16:0), (ii) sphingomyelin (d18:1/19:0, d19:1/18:0), (iii) sphingomyelin (d18:1/21:0, d17:1/22:0, d16:1/23:0), (iv) sphingomyelin (d18:1/25:0, d19:0/24:1, d20:1/23:0, d19:1/24:0), (v) sphingomyelin (d18:2/21:0, d16:2/23:0), (vi) sphingomyelin (d18:2/18:1), (vii) sphingomyelin (d17:2/16:0, d18:2/15:0), (viii) sphingomyelin (d17:1/16:0, d18:1/15:0, d16:1/17:0), (ix) sphingomyelin (d18:2/23:1), (x) sphingomyelin (d18:1/22:1, d18:2/22:0, d16:1/24:1), (xi) hydroxypalmitoyl sphingomyelin (d18:1/16:0(OH)). Additional associations are detailed in Table S3. See also Figure 1C for complementary associations in neurologically healthy controls.

### Figure S5.

A

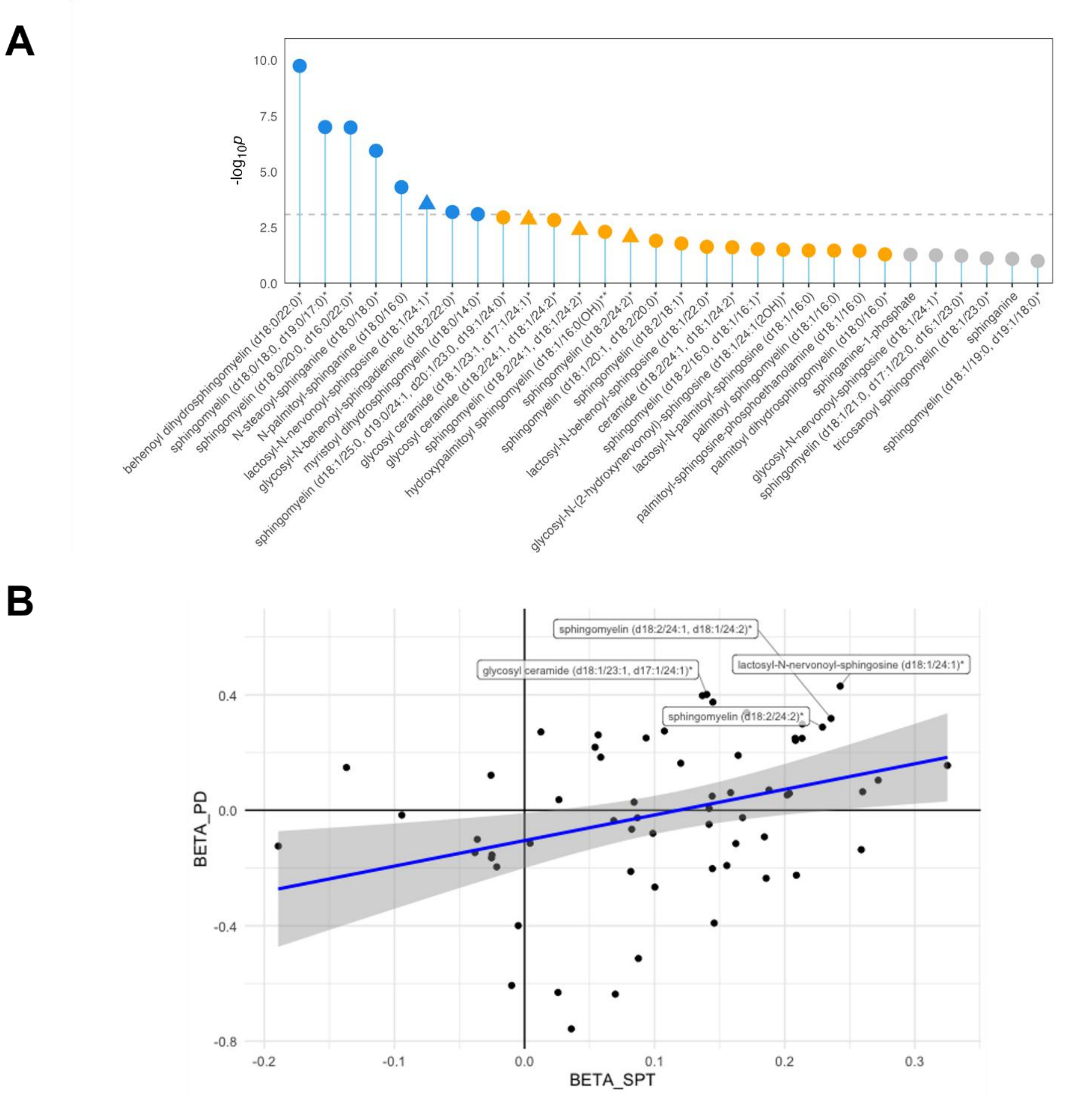


**Figure S5. Additional analyses of plasma sphingolipids in Parkinson's disease (PD) cases and control carriers of the *SPTSSB rs1450522* variant.** (A) Lollipop plots representing $-\log_{10}(p)$ from linear models comparing PD vs. Controls (corrected for age and sex), for top 30 associated metabolites. Blue dots are significant after multiple-testing correction, orange dots are suggestive associations (unadjusted $p < 0.05$), and gray dots are non-significant. Triangular lollipops represent metabolites associated with *SPTSSB rs1450522* among controls only (additive model, corrected for age and sex). (B) Scatterplot of metabolite associations with *SPTSSB rs1450522* in controls (x-axis, effect size, BETA_SPT) and PD versus controls (y-axis, effect size, BETA_PD). Analyses are adjusted for age and sex. Linear regression effect size = 0.87, p = 0.007. Labels highlight selected metabolites that are perturbed in PD cases and also associate with *SPTSSB rs1450522* in controls. See also Supplemental Table 3.

### Figure S6.

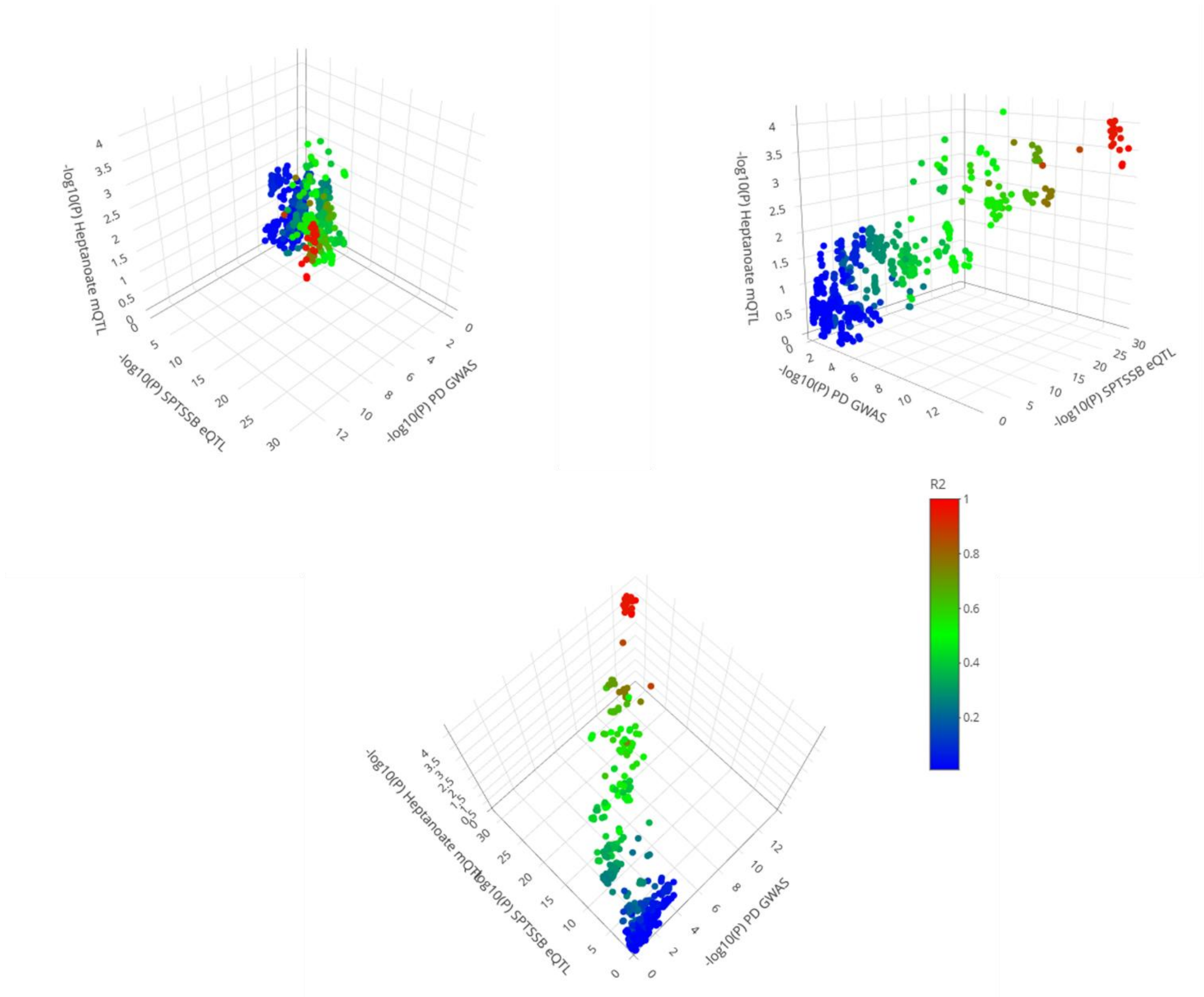


**Figure S6. Additional co-localization plots of Parkinson's disease (PD) risk variants at the SPTSSB locus with SPTSSB mRNA expression and fatty acid metabolism.** 3D scatterplot of *SPTSSB* variant associations with PD risk versus *SPTSSB* mRNA expression quantitative trait locus (eQTL) in excitatory neurons versus plasma heptanoate metabolite quantitative trait locus (mQTL). Color scale denotes linkage disequilibrium (LD, R2) with index variant, *rs1450522*.

### Figure S7.

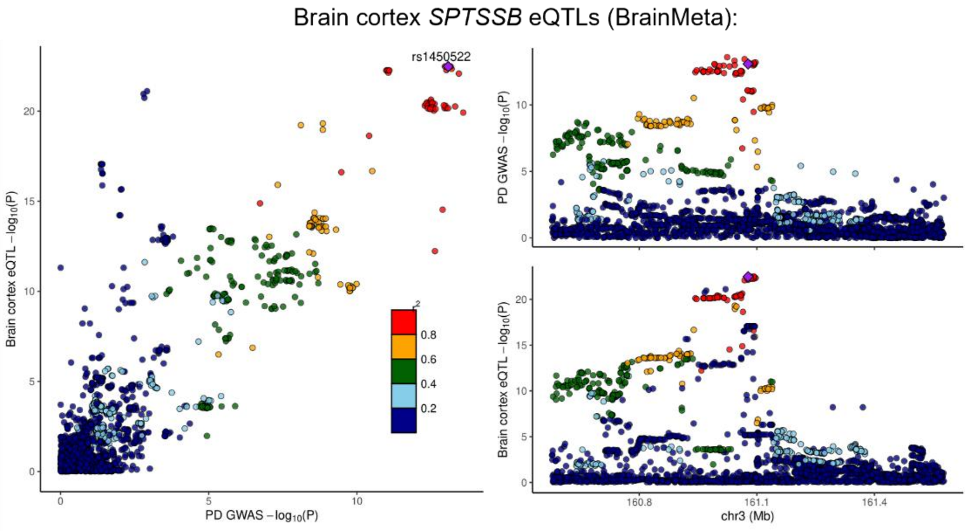


**Figure S7. Additional co-localization analysis of *SPTSSB* variant associations with Parkinson's disease (PD risk) versus *SPTSSB* mRNA expression in bulk cortex.** LocusCompare plots of of *SPTSSB* variant associations with PD risk versus *SPTSSB* mRNA expression quantitative trait locus (eQTL) in bulk cortical tissue from BrainMeta. Color scale denotes linkage disequilibrium (LD, R2) with index variant, *rs1450522*.

### Figure S8.

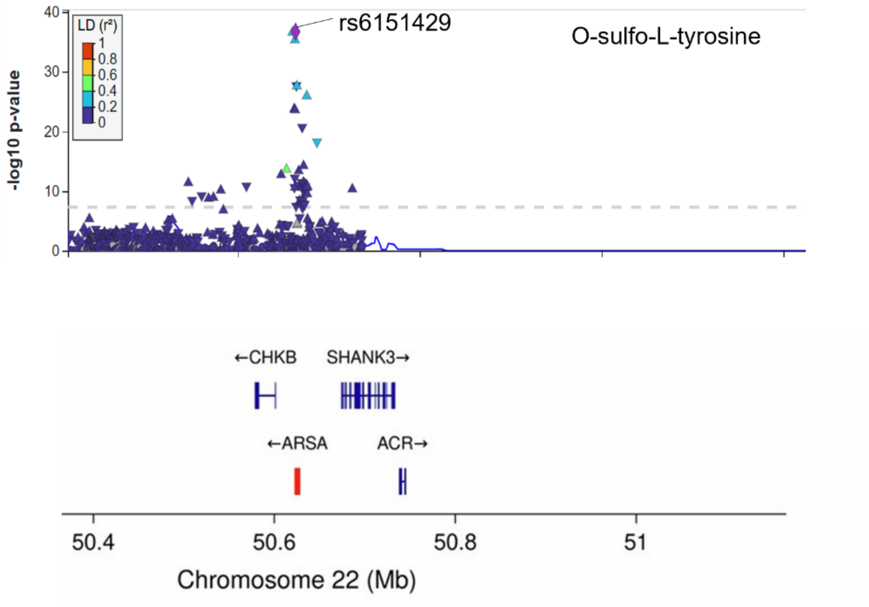


**Figure S8. Genetic variants at the *ARSA* locus are associated with O-sulfo-L-tyrosine plasma levels.** LocusCompare plot of *arylsulfatase A* (*ARSA*) metabolite quantitative trait locus (mQTL) in plasma. *rs6151429* is also associated with Parkinson's disease (PD) risk. Color scale denotes linkage disequilibrium (LD, R2) with index variant, rs6151429, which is significantly association with the metabolite O-sulfo-L-tyrosine ($p = 1.94 \times 10^{-37}$).

### Figure S9.

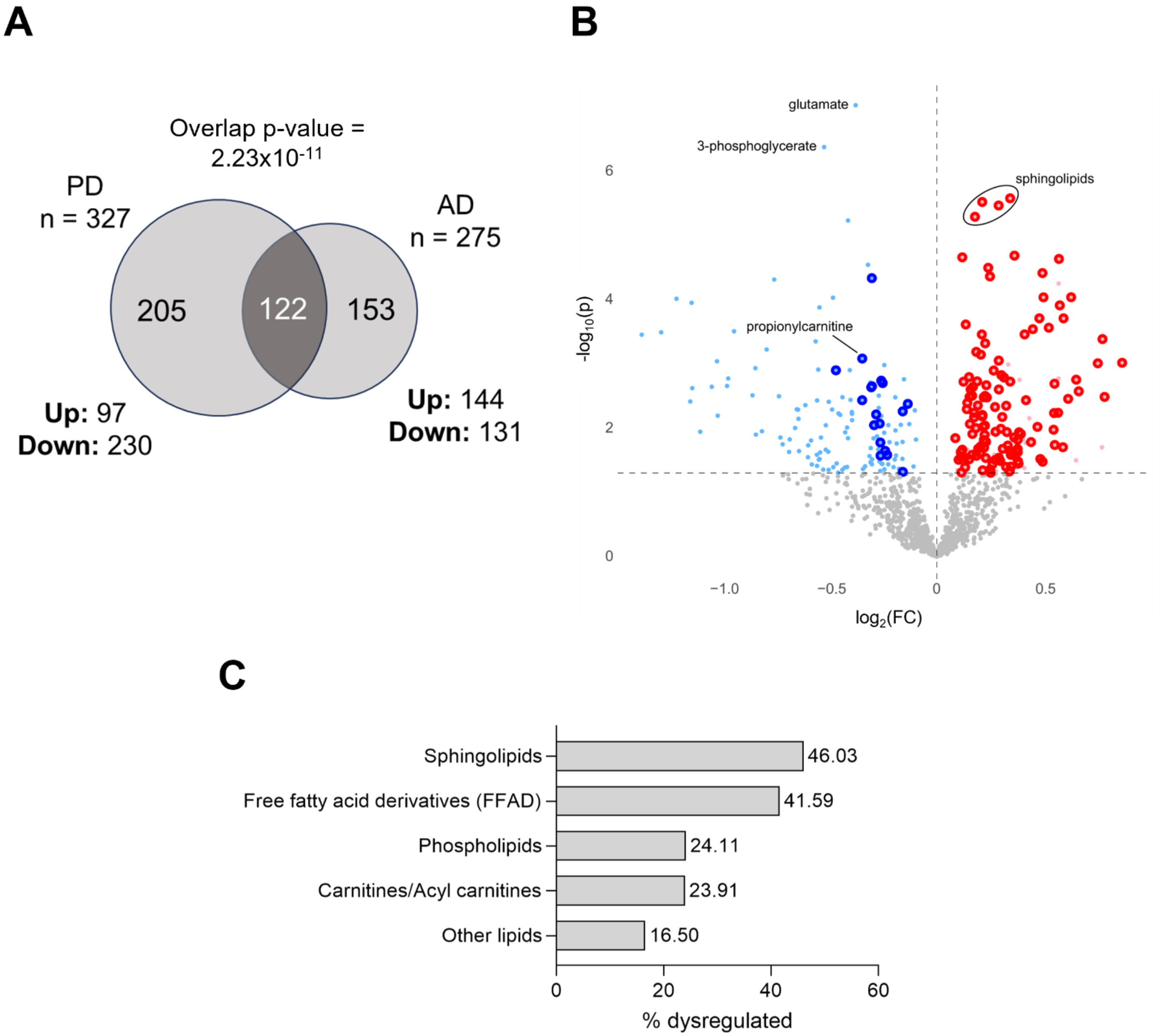


**Figure S9. Alzheimer's disease (AD) and Parkinson's disease (PD) reveal overlapping metabolic signatures from blood plasma.** Analyses corrected for age and sex. (A) Venn diagram showing unique and shared dysregulated metabolites from analyses of PD or AD subjects (versus controls). (B) Volcano plots show up- (red) or down-regulated (blue) meeting the suggestive significance threshold (unadjusted $p < 0.05$ dashed horizontal line). Lipid species are shown with larger, dark blue/red dots. (C) Plots showing the proportion (%) of dysregulated species within each lipid class. The sphingolipid and free fatty acid derivatives were significantly enriched for dysregulated metabolites, based on a Fisher test $p < 0.05$.

### Figure S10.

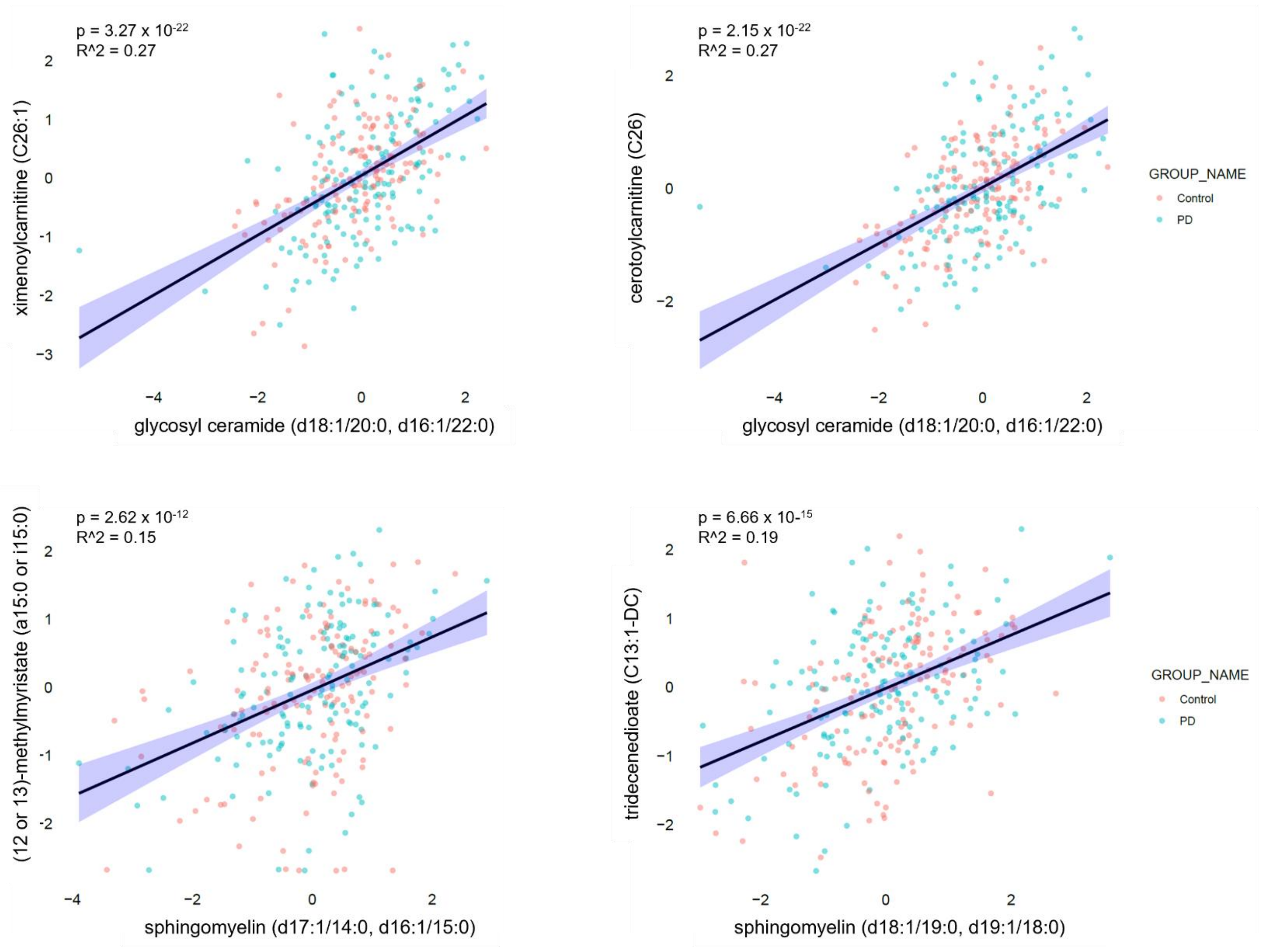


**Figure S10. Selected sphingolipids are correlated with acylcarnitine and fatty acid levels in blood plasma.** Scatterplots show sphingolipid metabolite levels (x-axis) versus acylcarnitine/fatty acid (y-axis). Plotted z-scores from linear regression analyses, corrected for age at sample collection, and sex. For all examples shown regression $p < 0.05$ and $R^2 > 0.15$. Colors denote Parkinson's disease cases (blue) and controls (red).

### Figure S11.

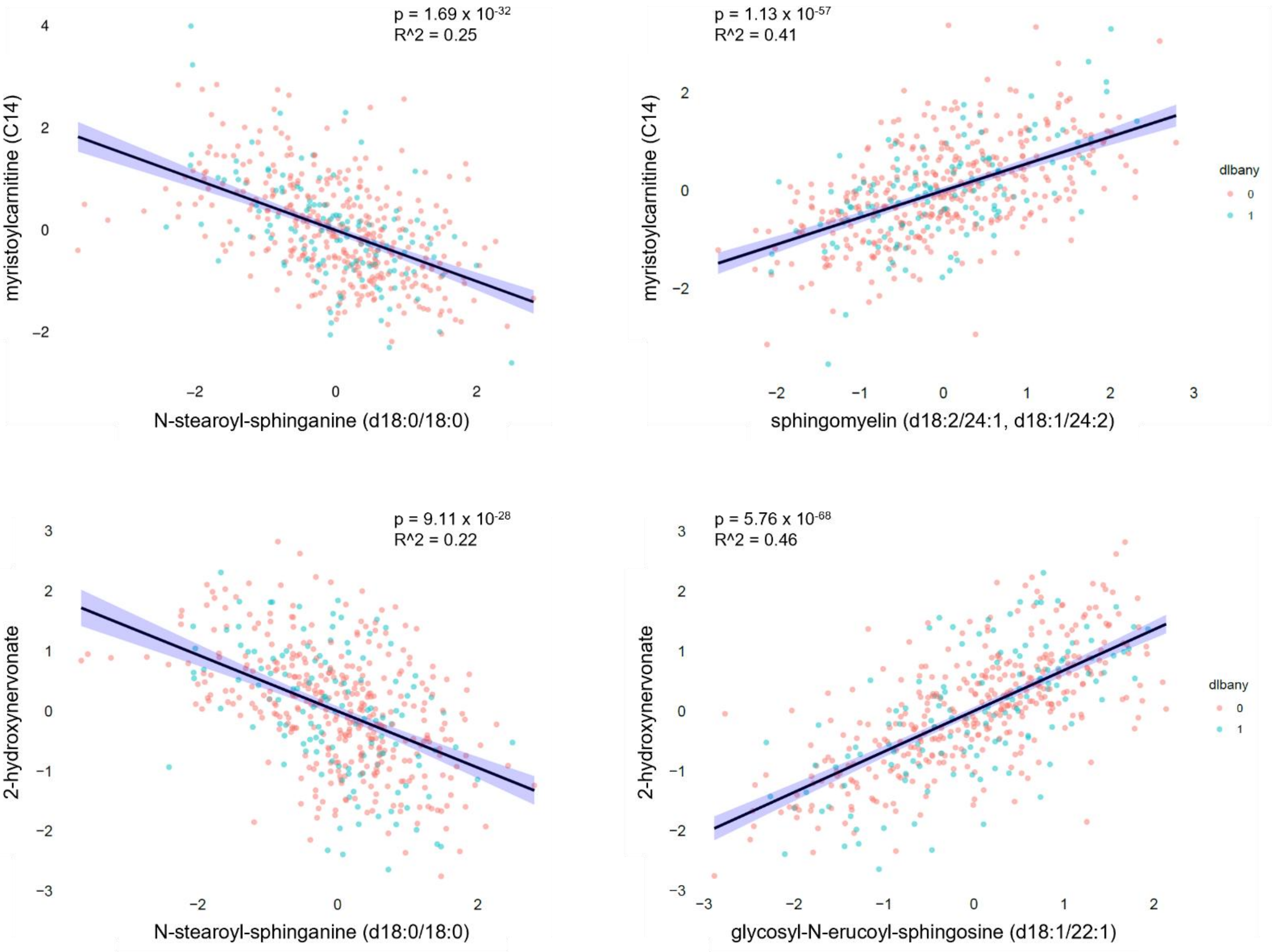


**Figure S11. Selected sphingolipids are correlated with acylcarnitine and fatty acid levels in brain.** Scatterplots show sphingolipid metabolite levels (x-axis) versus acylcarnitine/fatty acid (y-axis). Plotted z-scores from linear regression analyses, corrected for age at death, sex, and postmortem interval. For all examples shown regression $p < 0.05$ and $R^2 > 0.15$. Colors denote Parkinson's disease cases (blue) and controls (red).

### Table S1.

**Clinical and demographic features of the Houston PD case-control cohort.**

| | PD | Control |
|---|---|---|
| ***Subjects, N*** | 149 | 150 |
| ***Age at sample collection, mean (range)*** | 65.29 (34 – 85) | 66.3 (30 – 86) |
| ***Sex, N female (%)*** | 55 (36.91%) | 100 (66.67%) |
| ***Disease duration, mean (range)*** | 8 (1-30) | - |
| ***Reported race/ethnicity, N European (%)*** | 118 (79.19%) | 121 (80.67%) |
| ***Family history of PD, N (%)*** | 59 (39.60%) | 31 (20.67%) |

### Table S2.

**Clinical and demographic features from Religious Orders Study & Rush Memory and Aging Project.**

| | |
|---|---|
| ***Subjects, N*** | 490 |
| ***Pathology, N Lewy bodies (%)*** | 129 (26.33%) |
| ***Clinical diagnosis, N Parkinson's (%)*** | 35 (7.14%) |
| ***Age of death, mean (range)*** | 90.57 (71 – 106) |
| ***Sex, N female (%)*** | 345 (70.41%) |
| ***Post-mortem interval, mean (range)*** | 8.13 (0.87 – 43.88) |

### Table S3.

**Sphingolipid dysmetabolism in PD and *rs1450522* carriers.**

| Pathway | Sphingolipid | PD versus Control[1] Fold-change | PD versus Control[1] p-value | *rs1450522* (Controls)[2] Fold-change | *rs1450522* (Controls)[2] p-value | *rs1450522* (PD)[2] Fold-change | *rs1450522* (PD)[2] p-value |
|---|---|---|---|---|---|---|---|
| Ceramide PEs | palmitoyl-sphingosine-phosphoethanolamine (d18:1/16:0) | 1.062 | **0.023** | 1.049 | 0.120 | 1.025 | 0.377 |
| Ceramides | ceramide (d18:1/20:0, d16:1/22:0, d20:1/18:0) | 0.988 | 0.758 | 1.022 | 0.600 | 0.820* | **0.047*** |
| Ceramides | ceramide (d18:2/24:1, d18:1/24:2) | 1.107 | **0.018** | 1.005 | 0.914 | 1.049 | 0.361 |
| Dihydroceramides | N-palmitoyl-sphinganine (d18:0/16:0) | 0.794 | **$2.11 \times 10^{-5}$** | 1.039 | 0.553 | 1.077 | 0.181 |
| Dihydroceramides | N-stearoyl-sphinganine (d18:0/18:0) | 0.575 | **$2.03 \times 10^{-7}$** | 0.991 | 0.941 | 1.236 | 0.068 |
| Dihydrosphingomyelins | behenoyl dihydrosphingomyelin (d18:0/22:0) | 0.707 | **$2.80 \times 10^{-11}$** | 1.016 | 0.780 | 1.098 | 0.091 |
| Dihydrosphingomyelins | myristoyl dihydrosphingomyelin (d18:0/14:0) | 0.861 | **0.001** | 1.058 | 0.220 | 1.023 | 0.659 |
| Dihydrosphingomyelins | palmitoyl dihydrosphingomyelin (d18:0/16:0) | 0.938 | **0.037** | 1.050 | 0.157 | 1.017 | 0.632 |
| Dihydrosphingomyelins | sphingomyelin (d18:0/18:0, d19:0/17:0) | 0.707 | **$2.60 \times 10^{-8}$** | 1.014 | 0.849 | 1.130 | 0.059 |
| Dihydrosphingomyelins | sphingomyelin (d18:0/20:0, d16:0/22:0) | 0.705 | **$2.41 \times 10^{-8}$** | 1.038 | 0.596 | 1.133 | 0.061 |
| Hexosylceramides (HCER) | glycosyl ceramide (d18:1/23:1, d17:1/24:1) | 1.376 | **0.001** | 1.828* | **0.030*** | 1.140 | 0.216 |
| Hexosylceramides (HCER) | glycosyl ceramide (d18:2/24:1, d18:1/24:2) | 1.165 | **0.001** | 1.062 | 0.212 | 1.072 | 0.224 |
| Hexosylceramides (HCER) | glycosyl-N-(2-hydroxynervonoyl)-sphingosine (d18:1/24:1(2OH)) | 0.834 | **0.019** | 1.066 | 0.425 | 1.015 | 0.869 |
| Hexosylceramides (HCER) | glycosyl-N-behenoyl-sphingadienine (d18:2/22:0) | 1.187 | **0.001** | 1.062 | 0.237 | 1.101 | 0.109 |
| Lactosylceramides (LCER) | lactosyl-N-behenoyl-sphingosine (d18:1/22:0) | 1.123 | **0.022** | 1.026 | 0.672 | 0.998 | 0.976 |
| Lactosylceramides (LCER) | lactosyl-N-nervonoyl-sphingosine (d18:1/24:1) | 1.151 | **$2.74 \times 10^{-4}$** | 1.083 | **0.047** | 1.027 | 0.556 |
| Lactosylceramides (LCER) | lactosyl-N-palmitoyl-sphingosine (d18:1/16:0) | 1.074 | **0.029** | 1.060 | 0.108 | 1.027 | 0.455 |
| Sphingolipid Synthesis | sphinganine-1-phosphate | 0.917 | **0.040** | 1.033 | 0.498 | 0.988 | 0.805 |
| Sphingomyelins | hydroxypalmitoyl sphingomyelin (d18:1/16:0(OH)) | 1.101 | **0.005** | 1.050 | 0.228 | 1.182* | **0.035*** |
| Sphingomyelins | palmitoyl sphingomyelin (d18:1/16:0) | 1.039 | **0.026** | 1.014 | 0.487 | 1.017 | 0.368 |
| Sphingomyelins | sphingomyelin (d17:1/16:0, d18:1/15:0, d16:1/17:0) | 0.988 | 0.716 | 1.040 | 0.273 | 1.085 | **0.023** |
| Sphingomyelins | sphingomyelin (d17:2/16:0, d18:2/15:0) | 0.976 | 0.568 | 1.032 | 0.499 | 1.116 | **0.022** |
| Sphingomyelins | sphingomyelin (d18:1/17:0, d17:1/18:0, d19:1/16:0) | 1.011 | 0.707 | 1.038 | 0.272 | 1.108 | **0.002** |
| Sphingomyelins | sphingomyelin (d18:1/18:1, d18:2/18:0) | 1.042 | 0.124 | 1.029 | 0.312 | 1.064 | **0.045** |
| Sphingomyelins | sphingomyelin (d18:1/19:0, d19:1/18:0) | 0.934 | 0.078 | 1.054 | 0.218 | 1.133 | **0.003** |
| Sphingomyelins | sphingomyelin (d18:1/20:0, d16:1/22:0) | 0.970 | 0.203 | 1.055 | **0.044** | 1.065 | **0.019** |
| Sphingomyelins | sphingomyelin (d18:1/20:1, d18:2/20:0) | 1.072 | **0.009** | 1.050 | 0.081 | 1.066 | **0.040** |
| Sphingomyelins | sphingomyelin (d18:1/21:0, d17:1/22:0, d16:1/23:0) | 0.930 | **0.048** | 1.069 | 0.110 | 1.106 | **0.012** |
| Sphingomyelins | sphingomyelin (d18:1/22:1, d18:2/22:0, d16:1/24:1) | 1.032 | 0.143 | 1.068 | **0.005** | 1.055 | **0.035** |
| Sphingomyelins | sphingomyelin (d18:1/25:0, d19:0/24:1, d20:1/23:0, d19:1/24:0) | 0.835 | **$3.62 \times 10^{-4}$** | 0.998 | 0.970 | 1.154 | **0.013** |
| Sphingomyelins | sphingomyelin (d18:2/16:0, d18:1/16:1) | 1.056 | **0.020** | 1.046 | 0.080 | 1.040 | 0.144 |
| Sphingomyelins | sphingomyelin (d18:2/18:1) | 1.090 | **0.010** | 1.032 | 0.374 | 1.092 | **0.020** |
| Sphingomyelins | sphingomyelin (d18:2/21:0, d16:2/23:0) | 1.026 | 0.511 | 1.098 | **0.022** | 1.110 | **0.019** |
| Sphingomyelins | sphingomyelin (d18:2/23:0, d18:1/23:1, d17:1/24:1) | 1.030 | 0.345 | 1.082 | **0.020** | 1.072 | 0.055 |
| Sphingomyelins | sphingomyelin (d18:2/23:1) | 1.021 | 0.606 | 1.080 | 0.070 | 1.113 | **0.025** |
| Sphingomyelins | sphingomyelin (d18:2/24:1, d18:1/24:2) | 1.077 | **0.004** | 1.057 | **0.037** | 1.029 | 0.347 |
| Sphingomyelins | sphingomyelin (d18:2/24:2) | 1.102 | **0.006** | 1.078 | **0.049** | 1.034 | 0.409 |
| Sphingomyelins | stearoyl sphingomyelin (d18:1/18:0) | 0.995 | 0.843 | 1.021 | 0.496 | 1.076 | **0.014** |

1 Metabolite perturbations were compared between 149 PD cases and 150 controls without PD. Significance is based on the likelihood ratio test with adjustment for age and sex. Suggestive significance (bold) based on unadjusted $p < 0.05$.

2 Metabolite perturbations were compared based on *rs1450522* genotype, including noncarriers, heterozygotes, and homozygotes, with separate analyses conducted in PD cases and controls without PD. Regression models, with adjustment for age and sex, tested both an additive and recessive genetic model; asterisk (*) denotes results from the recessive model meeting the suggestive significance level ($p < 0.05$).

### Table S3.

**Associations of *SPTSSB* rs1450522$^{G}$ with gene expression in different cell types.**

| Cell type | Beta | Std. Error | P-value | FDR significant? |
|---|---|---|---|---|
| Astrocytes | -0.008 | 0.070 | 0.910 | No |
| Excitatory neurons | 0.391 | 0.030 | $8.14 \times 10^{-32}$ | Yes |
| Inhibitory neurons | 0.152 | 0.053 | $4.46 \times 10^{-3}$ | No |
| Oligodendrocytes | 0.015 | 0.071 | 0.832 | No |

No expression quantitative trait loci (eQTLs) were detected for endothelial cells, microglia, and oligodendrocyte precursor cells. All analyses were conducted with single-nucleus expression data from the Religious Orders Study and Rush Memory and Aging Project (ROSMAP).

### Table S5.

**Additional co-localization analysis of Parkinson's disease (PD) risk variants at the *SPTSSB* locus with candidate gene expression and fatty acid metabolism.**

| Phenotype 1 | Phenotype 2 | Gene expression | N of SNPs | PP.H0.abf | PP.H1.abf | PP.H2.abf | PP.H3.abf | PP.H4.abf |
|---|---|---|---|---|---|---|---|---|
| PD | Brain cortex eQTLs | *PPM1L* | none | - | - | - | - | - |
| PD | Brain cortex eQTLs | *B3GALNT1* | none | - | - | - | - | - |
| PD | Brain cortex eQTLs | *NMD3* | 2,797 | 0.000 | 0.000 | 0.000 | 0.123 | 0.877 |
| PD | Brain cortex eQTLs | *SPTSSB* | 2,797 | 0.000 | 0.000 | 0.000 | 0.037 | 0.963 |
| PD | Brain cortex eQTLs | *OTOL1* | none | - | - | - | - | - |
| PD | Whole blood eQTLs | *PPM1L* | 2,180 | 0.000 | 0.785 | 0.000 | 0.194 | 0.020 |
| PD | Whole blood eQTLs | *B3GALNT1* | 2,511 | 0.000 | 0.641 | 0.000 | 0.190 | 0.169 |
| PD | Whole blood eQTLs | *NMD3* | 2,511 | 0.000 | 0.047 | 0.000 | 0.072 | 0.881 |
| PD | Whole blood eQTLs | *SPTSSB* | 2,511 | 0.000 | 0.862 | 0.000 | 0.118 | 0.020 |
| PD | Whole blood eQTLs | *OTOL1* | none | - | - | - | - | - |
| PD | Excitatory neurons eQTLs | *PPM1L* | 1,565 | 0.000 | 0.778 | 0.000 | 0.185 | 0.038 |
| PD | Excitatory neurons eQTLs | *B3GALNT1* | 1,847 | 0.000 | 0.878 | 0.000 | 0.098 | 0.023 |
| PD | Excitatory neurons eQTLs | *NMD3* | 1,847 | 0.000 | 0.000 | 0.000 | 1.000 | 0.000 |
| PD | Excitatory neurons eQTLs | *SPTSSB* | 1,847 | 0.000 | 0.000 | 0.000 | 0.034 | 0.966 |
| PD | Excitatory neurons eQTLs | *OTOL1* | none | - | - | - | - | - |
| PD | Plasma heptanoate mQTLs | - | 1,481 | 0.000 | 0.052 | 0.000 | 0.054 | 0.894 |
| Excitatory neurons eQTLs | Plasma heptanoate mQTLs | *SPTSSB* | 911 | 0.000 | 0.051 | 0.000 | 0.049 | 0.900 |

### Table S6.

**Mitochondrial pathways from Lewy body-associated differential protein expression.**

| KEGG Pathway | Protein symbol | Ortholog ID | LRT P-value | Fold-change |
|---|---|---|---|---|
| Fatty acid degradation (hsa00071) | ACAA2 | KO:K07508 | 0.037 | 0.979 |
| | ACAT2 | KO:K00626 | 0.001 | 1.032 |
| | ACOX3 | KO:K00232 | 0.009 | 1.031 |
| | ACSL3 | KO:K01897 | 0.022 | 0.988 |
| | ALDH2 | KO:K00128 | 0.021 | 0.977 |
| | CPT1A | KO:K08765 | 0.014 | 0.978 |
| | CPT1C | KO:K19524 | 0.021 | 1.020 |
| | ECI1 | KO:K13238 | 0.012 | 0.966 |
| | GCDH | KO:K00252 | 0.049 | 0.973 |
| OXPHOS (hsa00190) | ATP5J | KO:K02131 | 0.010 | 0.976 |
| | ATP6V1F | KO:K02151 | 0.015 | 0.983 |
| | COX4I1 | KO:K02263 | 0.005 | 0.974 |
| | COX5B | KO:K02265 | 0.008 | 0.977 |
| | NDUFA10 | KO:K03954 | 0.036 | 0.979 |
| | NDUFA8 | KO:K03952 | 0.035 | 0.978 |
| | NDUFB3 | KO:K03959 | 0.034 | 0.975 |
| | NDUFB5 | KO:K03961 | 0.046 | 0.982 |
| | NDUFB7 | KO:K03963 | 0.030 | 0.972 |
| | NDUFB9 | KO:K03965 | 0.049 | 0.980 |
| | NDUFC2 | KO:K03968 | 0.040 | 0.979 |
| | NDUFS6 | KO:K03939 | 0.033 | 0.979 |
| | PPA2 | KO:K01507 | 0.024 | 0.981 |
| | UQCRB | KO:K00417 | 0.048 | 0.985 |
| | UQCRC1 | KO:K00414 | 0.027 | 0.985 |
| | UQCRQ | KO:K00418 | 0.040 | 0.983 |
| TCA cycle (hsa00020) | DLAT | KO:K00627 | 0.021 | 0.981 |
| | DLD | KO:K00382 | 0.050 | 0.988 |
| | IDH2 | KO:K00031 | 0.003 | 0.971 |
| | PDHA1 | KO:K00161 | 0.001 | 0.976 |
| | PDHB | KO:K00162 | 0.006 | 0.975 |
| | SUCLG1 | KO:K01899 | 0.015 | 0.982 |
